# Dissecting the genetic complexity of myalgic encephalomyelitis/chronic fatigue syndrome via deep learning-powered genome analysis

**DOI:** 10.1101/2025.04.15.25325899

**Authors:** Sai Zhang, Fereshteh Jahanbani, Varuna Chander, Martin Kjellberg, Menghui Liu, Katherine A. Glass, David S. Iu, Faraz Ahmed, Han Li, Rajan Douglas Maynard, Tristan Chou, Johnathan Cooper-Knock, Martin Jinye Zhang, Durga Thota, Michael Zeineh, Jennifer K. Grenier, Andrew Grimson, Maureen R. Hanson, Michael P. Snyder

**Affiliations:** Department of Genetics, Center for Genomics and Personalized Medicine, Stanford University School of Medicine, Stanford, CA, USA; Department of Epidemiology, University of Florida, Gainesville, FL, USA; Department of Biostatistics & Biomedical Engineering, Genetics Institute, McKnight Brain Institute, University of Florida, Gainesville, FL, USA; Department of Molecular Biology and Genetics, Cornell University, Ithaca, NY, USA; Genomics Facility, Biotechnology Resource Center, Cornell University, Ithaca, NY, USA; School of Mathematical Sciences and LPMC, Nankai University, Tianjin, China; Sheffield Institute for Translational Neuroscience, University of Sheffield, Sheffield, UK; Ray and Stephanie Lane Computational Biology Department, Carnegie Mellon University, Pittsburgh, PA, USA; Department of Radiology, Stanford University School of Medicine, Stanford, CA, USA

## Abstract

Myalgic encephalomyelitis/chronic fatigue syndrome (ME/CFS) is a complex, heterogeneous, and systemic disease defined by a suite of symptoms, including unexplained persistent fatigue, post-exertional malaise (PEM), cognitive impairment, myalgia, orthostatic intolerance, and unrefreshing sleep. The disease mechanism of ME/CFS is unknown, with no effective curative treatments. In this study, we present a multi-site ME/CFS whole-genome analysis, which is powered by a novel deep learning framework, HEAL2. We show that HEAL2 not only has predictive value for ME/CFS based on personal rare variants, but also links genetic risk to various ME/CFS-associated symptoms. Model interpretation of HEAL2 identifies 115 ME/CFS-risk genes that exhibit significant intolerance to loss-of-function (LoF) mutations. Transcriptome and network analyses highlight the functional importance of these genes across a wide range of tissues and cell types, including the central nervous system (CNS) and immune cells. Patient-derived multi-omics data implicate reduced expression of ME/CFS risk genes within ME/CFS patients, including in the plasma proteome, and the transcriptomes of B and T cells, especially cytotoxic CD4 T cells, supporting their disease relevance. Pan-phenotype analysis of ME/CFS genes further reveals the genetic correlation between ME/CFS and other complex diseases and traits, including depression and long COVID-19. Overall, HEAL2 provides a candidate genetic-based diagnostic tool for ME/CFS, and our findings contribute to a comprehensive understanding of the genetic, molecular, and cellular basis of ME/CFS, yielding novel insights into therapeutic targets. Our deep learning model also offers a potent, broadly applicable framework for parallel rare variant analysis and genetic prediction for other complex diseases and traits.

## Introduction

Myalgic encephalomyelitis/chronic fatigue syndrome (ME/CFS) is a complex and debilitating multisystem disorder that affects millions of people, with an estimated 65 million individuals living with the condition worldwide prior to 2020^1^. It has a complex and heterogeneous manifestation that presents with a broad spectrum of debilitating symptoms. ME/CFS occurs with other debilitating symptoms, such as post-exertional malaise, unrefreshing sleep, and cognitive dysfunction^2^. The impact of this condition is far-reaching, with one in four patients becoming housebound or bedbound, and many of the most severely affected individuals requiring feeding tubes^3^. As a significant diagnostic challenge, ME/CFS is often misdiagnosed or undiagnosed due to the lack of a definitive biomarker, with 90% of the individuals frequently undergoing a prolonged diagnostic odyssey after years of medical consultation^4^.

ME/CFS can affect individuals of all ages, races, ethnicities, and socioeconomic backgrounds, making it a universal threat^5^. Unfortunately, there are no FDA-approved treatments for the disease, and current management strategies focus primarily on symptom relief. This lack of specific treatments highlights the urgent need for more robust research into the disease’s etiology and pathogenesis.

Despite its significant impact, ME/CFS remains poorly understood, with its underlying mechanisms still largely unknown. A combination of genetic, infectious, and environmental factors may contribute to disease onset and progression. A majority of individuals with ME/CFS report a preceding viral-like infection and clustered outbreaks have occurred in the past^6^. However, not everyone who suffers a viral infection develops ME/CFS nor can all cases be traced to a symptomatic viral illness; therefore understanding the genetic basis of susceptibility to ME/CFS is critical, as it could not only illuminate the underlying mechanisms of the disease, but also open the door for novel diagnostics and therapeutic interventions. Recent advances in genomics and the growing recognition that ME/CFS exhibits genetic risk factors^7^ present new opportunities for research. Previous genetic studies^8,9^ have been limited by small sample sizes and inconsistent results, highlighting the need for large-scale, well-characterized patient cohorts, and advanced methodology to uncover meaningful genetic insights.

We provide a large whole-genome analysis of ME/CFS, powered by the novel deep learning framework HEAL2. Based on rare coding variants, HEAL2 predicted ME/CFS risk for individuals across multiple cohorts. This approach further identified 115 risk genes associated with ME/CFS exhibiting critical functional roles across various tissues and cell types, including the central nervous system (CNS) and immune cells. Multi-omics data from ME/CFS patients validated the relevance of these findings, showing reduced expression of these genes in affected individuals. Finally, our pan-phenotype analysis uncovered a genetic correlation between ME/CFS and other conditions, such as depression and long COVID-19. Our study not only contributes to a deeper understanding of the genetic basis of ME/CFS, but also provides a powerful, generic framework for conducting rare variant analysis in other complex diseases. By deriving a genetic risk score and identifying some genetic risk factors associated with ME/CFS, this research holds the future potential to catalyze the development of precision diagnosis and effective therapies, providing much-needed hope for the millions of people living with this life-altering disease.

## Results

### Whole-genome sequencing of multiple ME/CFS cohorts

We conducted whole-genome sequencing (WGS) on the same platform for *N* = 1,075 individuals (*N* = 464 ME/CFS patients, *N* = 611 negative controls) from three ME/CFS cohorts (**Fig. 1A**; **Methods**): (1) a Stanford cohort that we assembled (*N* = 208 cases, *N* = 534 controls; see methods), (2) a UK CureME cohort (*N* = 190 cases, *N* = 30 controls), and (3) a Cornell cohort (*N* = 66 cases, *N* = 47 controls). To increase the sample size of ME/CFS cases, we combined the Stanford and CureME cohorts into the discovery cohort and used the Cornell cohort as an independent testing cohort. After read alignment, variant calling, stringent quality controls (QCs), and ancestry analysis (**Methods**), we obtained the analysis-ready discovery cohort (*N* = 247 cases, *N* = 192 controls) and testing cohort (*N* = 36 cases, *N* = 21 controls). In particular, non-European and genetically-related individuals were excluded to control sampling bias (**Methods**).

**Figure 1.**
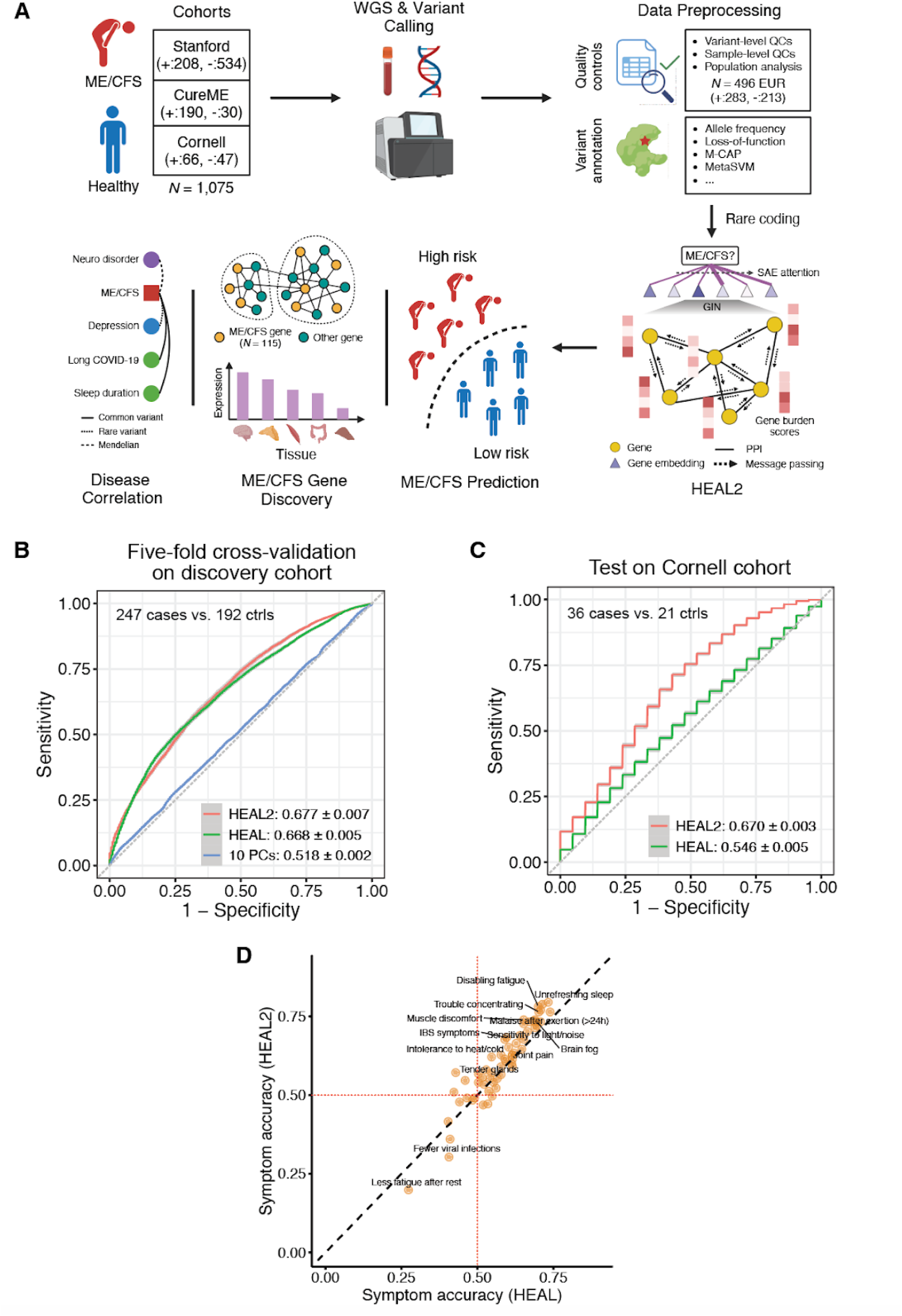
HEAL2 study design and prediction performance. (A) Schematic of our study design and HEAL2 model architecture. +, ME/CFS case; –, negative control; WGS, whole-genome sequencing; QCs, quality controls; EUR, European; GIN, graph isomorphism network; SAE, sparse autoencoder; PPI, protein-protein interaction. (B) Prediction performance of five-fold cross-validation (100 repeats) on the discovery cohort. The curve and shaded area represent the AUROC mean and 95% confidence interval (CI), respectively. AUROC, area under the receiver operating characteristic curve; PC, principal component. (C) Prediction performance of testing (500 repeats) on the independent Cornell cohort. The curve and shaded area represent the AUROC mean and 95% CI, respectively. (D) Correlation between genetic risk scores and patient symptoms. We examined the correlation by computing the accuracy of HEAL or HEAL2 on predicting different patient symptoms. The red dashed lines indicate the accuracy of 0.5 which is a random guess. IBS, irritable bowel syndrome.

We performed variant annotation for all genetic variants, including the single nucleotide variants (SNVs) and small insertions and deletions (indels), within the analysis-ready cohorts (**Fig. 1A**; **Methods**). In this study, we mainly focused on the coding variants including the loss-of-function (LoF) variants and missense (nonsynonymous) variants, wherein missense variants were further annotated using 38 functional scores (**Methods**). We have focused on rare variants, which are more likely to be functionally significant^10^, and are less likely to be confounded by linkage disequilibrium (LD). Given the moderate sample size of our cohorts, we used gnomAD^11^ (version 4.1, Non-Finnish European [NFE]) to estimate allele frequencies (AFs), and retained 99,958 rare coding variants (NFE AF < 1%) for downstream machine learning analysis (**Fig. 1A**).

### HEAL2 predicts ME/CFS risk from personal rare variants

To model the rare-variant genetic architecture of ME/CFS, we developed HEAL2, a graph neural network (GNN)-based framework that predicts ME/CFS risk from personal rare coding variants (**Fig. 1A**; **Methods**). HEAL2 extends our previous method HEAL^12^ in three aspects: (1) HEAL2 incorporates more comprehensive variant categories and functional scores; (2) HEAL2 employs an attention mechanism to improve model interpretation; (3) HEAL2 contains a non-linear GNN component based on the protein-protein interaction (PPI) network to capture the epistasis underpinning phenotypes, while HEAL is a linear model. Briefly speaking, for each individual HEAL2 first computes gene-level burden scores based on a variety of variant categories and functional scores using max and sum pooling (**Fig. 1A**; **Methods**). Next, HEAL2 conducts message passing of gene embeddings based on known PPIs. A sparse autoencoder (SAE)-based attention operation is then placed over gene embeddings after GNN to facilitate gene prioritization. Gene embeddings are pooled using attentions to construct a network embedding, which is used to compute the final ME/CFS risk score.

We first assessed the capacity of HEAL2 in modeling nonlinear genotype-phenotype (G2P) mapping using simulations. Based on a simulated nonlinear G2P dataset, we found that HEAL2 accurately predicted the disease from rare variants (area under the receiver operating characteristic curve [AUROC]: 0.891 [mean] ± 0.002 [95% CI]; area under the precision-recall curve [AUPRC]: 0.876 ± 0.001; **Supplementary** Fig. 1A and 1B), significantly outperforming HEAL in comparison (AUROC: 0.598 ± 0.004; AUPRC: 0.548 ± 0.005; **Supplementary** Fig. 1A and 1B). Similar results were observed on an independent dataset (AUROC: 0.886 ± 0.001 [HEAL2] vs. 0.528 ± 0.005 [HEAL]; AUPRC: 0.944 ± 0.001 [HEAL2] vs. 0.709 ± 0.004 [HEAL]; **Supplementary** Fig. 1C and 1D). These data confirmed the superiority of HEAL2 in genetic risk prediction in the context of nonlinear G2P.

To evaluate the performance of HEAL2 on real ME/CFS data, we first carried out a five-fold cross-validation (5-CV) based on the discovery cohort (**Methods**). Performance variance was estimated by repeating the 5-CV 100 times using different random seeds controlling dataset split and model initialization. We benchmarked HEAL2 against HEAL as our baseline using the same 5-CV procedure. Prediction performance was measured using the area under the receiver operating characteristic curve (AUROC) and the area under the precision-recall curve (AUPRC). Overall, HEAL2 exhibited better prediction performance (AUROC: 0.677 [mean] ± 0.007 [95% CI]; AUPRC: 0.727 ± 0.006; **Fig. 1B**, **Supplementary Fig. 2A**) than HEAL (AUROC: 0.668 ± 0.005; AUPRC: 0.716 ± 0.004; **Fig. 1B**, **Supplementary Fig. 2A**), suggesting that gene interactions might be a significant contributor to ME/CFS pathogenesis. Of note, a logistic regression model using the first 10 principal components (PCs) as its features yielded nearly random prediction (AUROC: 0.518 ± 0.002; AUPRC: 0.586 ± 0.002; **Fig. 1B, Supplementary Fig. 2A**), indicating the population homogeneity of our cohort after QCs.

Next, we evaluated HEAL2 based on the independent Cornell cohort wherein the model was trained on the discovery cohort (**Methods**). This train-test procedure was repeated 500 times with different random seeds. Notably, HEAL2 yielded comparable prediction performance (AUROC: 0.670 ± 0.003; AUPRC: 0.763 ± 0.002; **Fig. 1C**, **Supplementary Fig. 2B**) with that of 5-CV, and substantially outperformed HEAL (AUROC: 0.546 ± 0.005; AUPRC: 0.675 ± 0.004; **Fig. 1C**, **Supplementary Fig. 1B**). This result highlights the superiority of HEAL2 in generalizability over HEAL, indicating that HEAL2 captured biology-relevant genetic factors contributing to ME/CFS.

### HEAL2 risk score correlates with ME/CFS-relevant symptoms

Given the phenotypic heterogeneity of ME/CFS patients, we further asked whether our genetic risk score could precisely characterize diverse ME/CFS symptoms and severity. In particular, we linked HEAL and HEAL2 scores to various symptoms measured for ME/CFS patients within the CureME cohort (**Supplementary Table 1**; **Methods**). Surprisingly, we observed that although detailed phenotype data were not seen by the model, HEAL2 risk score was still strongly correlated with many symptoms showing different aspects of ME/CFS manifestation and disease severity (**Fig. 1D**), such as unrefreshing sleep, brain fog, malaise after exertion, and muscle discomfort. Of note, the HEAL2 score yielded sharper correlation than the HEAL score, implicating that HEAL2 better captured the phenotypic variation.

### HEAL2 identifies 115 ME/CFS risk genes

Model interpretation enables us to extract biological insights from the trained model. We first assessed the importance of different variant categories and scores in their contribution to prediction (**Methods**). Not unexpectedly, LoF variants, including stop gain/loss, start loss, and frameshift indels, contributed most to prediction (**Fig. 2A**), consistent with previous findings on the larger effect size of LoF variants compared to missense variants^13,14^. This was followed by evolution-based functional scores for missense variants (**Fig. 2A**), such as LINSIGHT^15^ and PrimateAI^16^.

**Figure 2.**
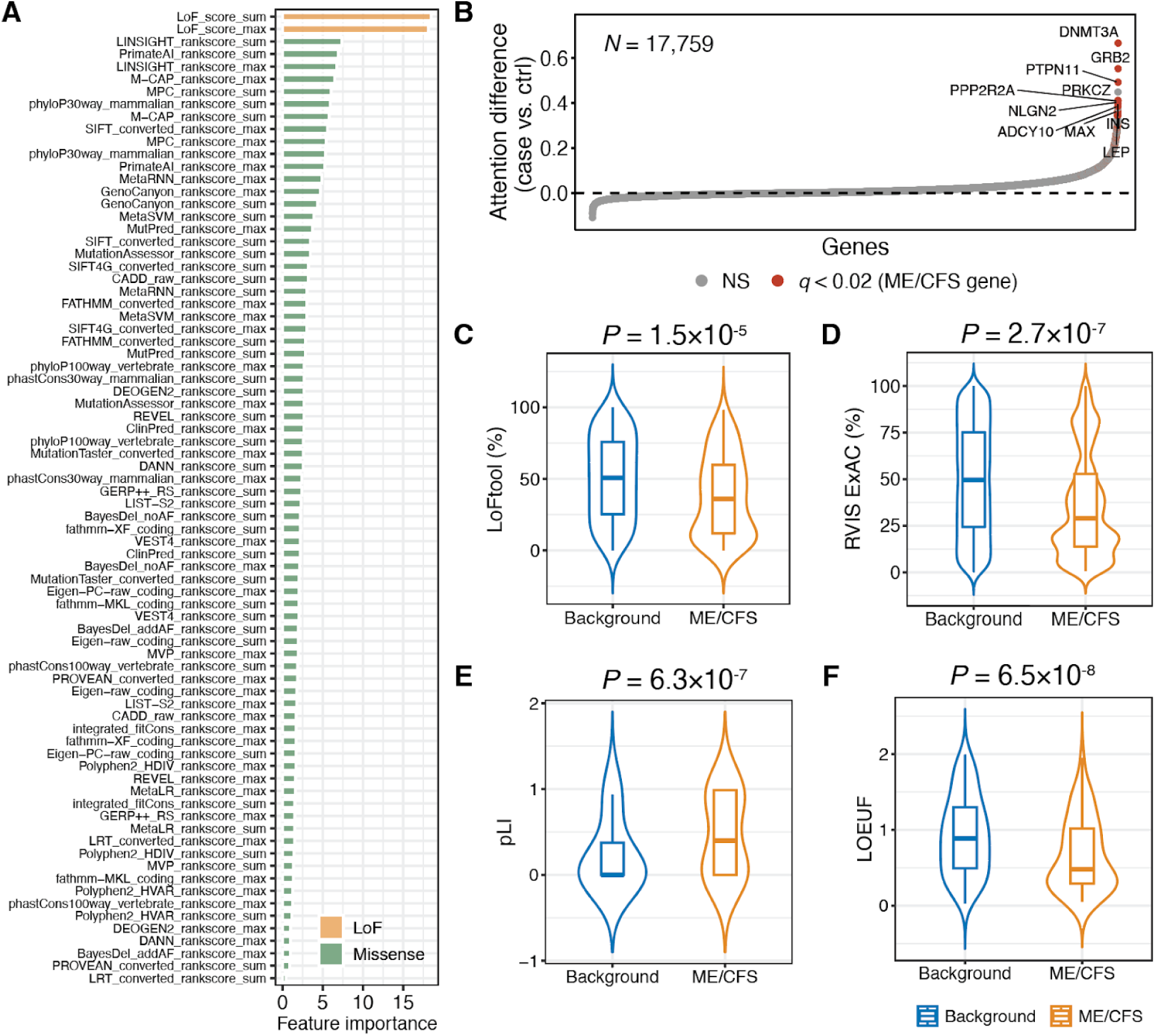
HEAL2 model interpretation and intolerance to LoF variants for ME/CFS genes. (A) HEAL2 input feature importance. LoF, loss-of-function. (B) Gene prioritization by HEAL2 based on attention scores. Ctrl, control; NS, not significant. *q*-value by the Storey-Tibshirani procedure. Top 10 significant genes were highlighted in red. (C-D) Intolerance to LoF variants for ME/CFS genes based on LoFtool (C), RIVS (D), pLI (E), and LOEUF (F). The box plot center line, limits, and whiskers represent the median, quartiles, and 1.5x interquartile range (IQR), respectively. pLI, probability of loss-of-function intolerance; LOEUF, loss-of-function observed/expected upper bound fraction. *P*-value by the two-sided Wilcoxon rank-sum test.

Gene-level attention scores derived from HEAL2 inform disease-gene associations. Using simulations, we first demonstrated that HEAL2 precisely prioritized disease-causing genes based on the gene-level attention (AUROC: 0.959; AUPRC: 0.505; **Supplementary** Fig. 1E and 1F; **Methods**). For ME/CFS, HEAL2 prioritized 115 genes that presented consistently larger attention scores among patients compared to controls (*q*-value < 0.02, Storey-Tibshirani procedure^17^; **Fig. 2B**, **Supplementary Table 2**; **Methods**). We defined these 115 genes as HEAL2-identified ME/CFS risk genes throughout this study. To gain insights into gene function, we first examined the sensitivity to LoF variants for HEAL2 ME/CFS genes based on four scores (**Methods**), including LoFtool^18^, RIVS^19^ (derived from the ExAC cohort^20^), pLI^20^, and LOEUF^21^. Interestingly, we found that ME/CFS risk genes exhibited significantly stronger intolerance to LoF variants compared to all protein-coding genes (*P* < 0.05, two-sided Wilcoxon rank-sum test; **Fig. 2C-2F**), suggesting their functional importance in shaping human health.

### ME/CFS genes display functional diversity across human tissues and cell types

To dissect the functional heterogeneity of HEAL2 ME/CFS genes, we next sought to systematically examine their expression patterns across different human tissues. By comparing expression levels between ME/CFS genes and the background transcriptome^22^ (**Methods**), we observed higher expression of ME/CFS genes spanning multiple tissues (adjusted *P* < 0.05, two-sided *t*-test with Bonferroni correction; **Fig. 3A**), including cerebral cortex, skeletal muscle, and colon. To obtain a finer-resolution, we further analyzed single-cell RNA-seq data^22^ (**Methods**) and revealed higher expression of ME/CFS genes across various cell types (adjusted *P* < 0.05, two-sided *t*-test with Bonferroni correction; **Fig. 3B**), including neurons, smooth muscle cells, and immune cells. At the protein level^23^ (**Methods**), we confirmed the higher expression of ME/CFS genes (*P* < 0.05, two-sided *t*-test; **Fig. 3C**) within the central nervous system (CNS). These results are consistent with tissues and organs affected by ME/CFS^24,25^, implicating their causal roles in impacting the disease risk and symptoms.

**Figure 3.**
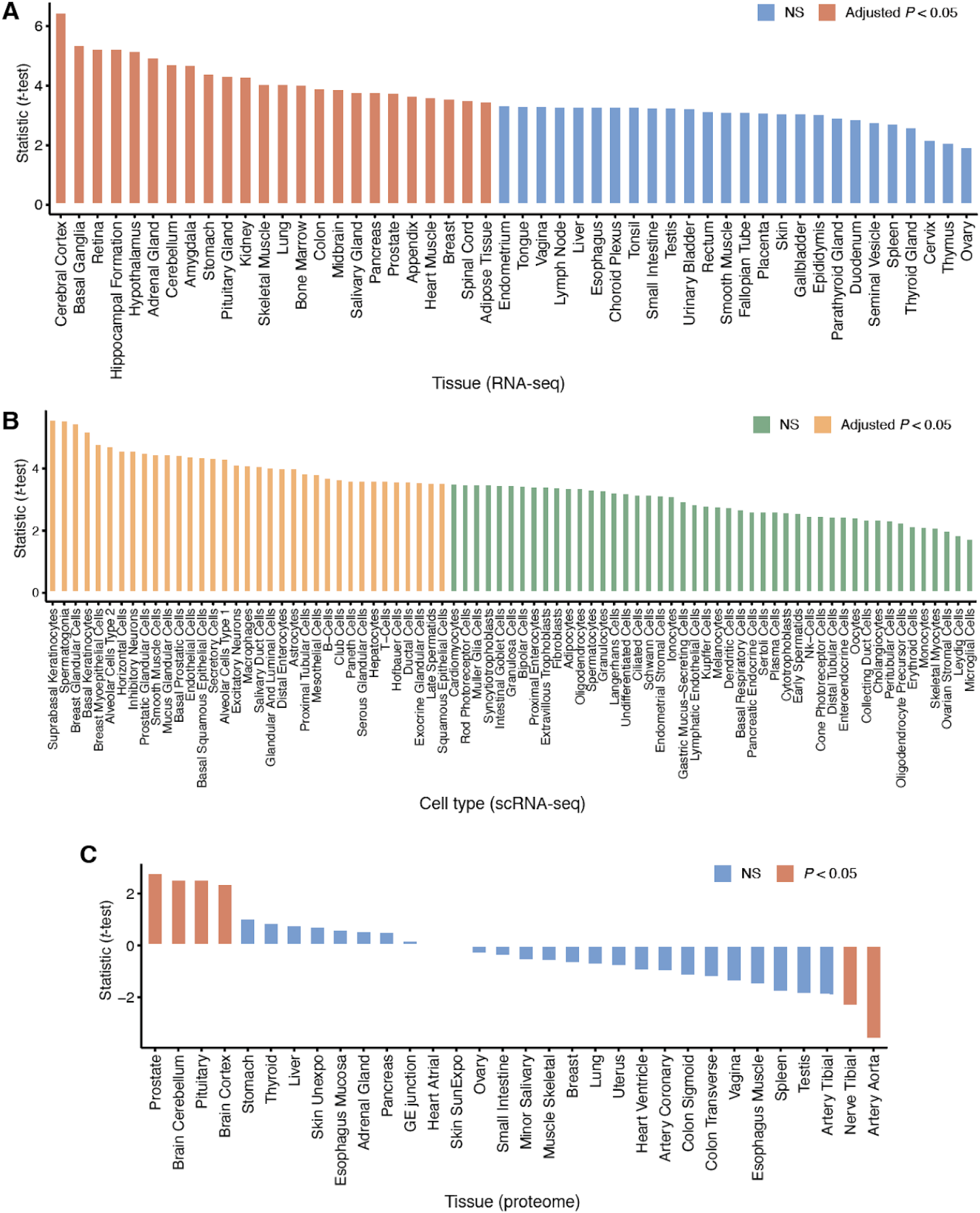
Expression patterns of ME/CFS genes across different human tissues and cell types. (A-B) Gene expression comparison between ME/CFS genes and the transcriptome within 50 human tissues (A) and 81 cell types (B). NS, not significant. *P*-value by two-sided *t*-test. Multiple testing correction was performed using the Bonferroni procedure. (C) Protein expression comparison between ME/CFS genes and all protein-coding genes within 32 human tissues. *P*-value by two-sided *t*-test.

To investigate the function of our ME/CFS genes at a systems level, we further carried out a network analysis^12,26–29^ (**Methods**) by mapping 115 ME/CFS genes onto a protein-protein interaction (PPI) network^30^. We assessed the enrichment of ME/CFS genes within different gene modules of the PPI network. Notably, four gene modules (out of 1,261 modules) were identified to be significantly enriched with ME/CFS genes (false discovery rate [FDR] < 0.05, Fisher’s exact test; **Fig. 4A** and **4B**, **Supplementary Fig. 3**). Gene ontology (GO) analysis showed that M9 genes were associated with proteasome function and particularly degradation of ubiquitinated proteins that are targeted for turnover (**Fig. 4C**), and M20 genes were linked to synaptic function (**Fig. 4D**). These findings reveal the functional diversity of ME/CFS risk genes.

**Figure 4.**
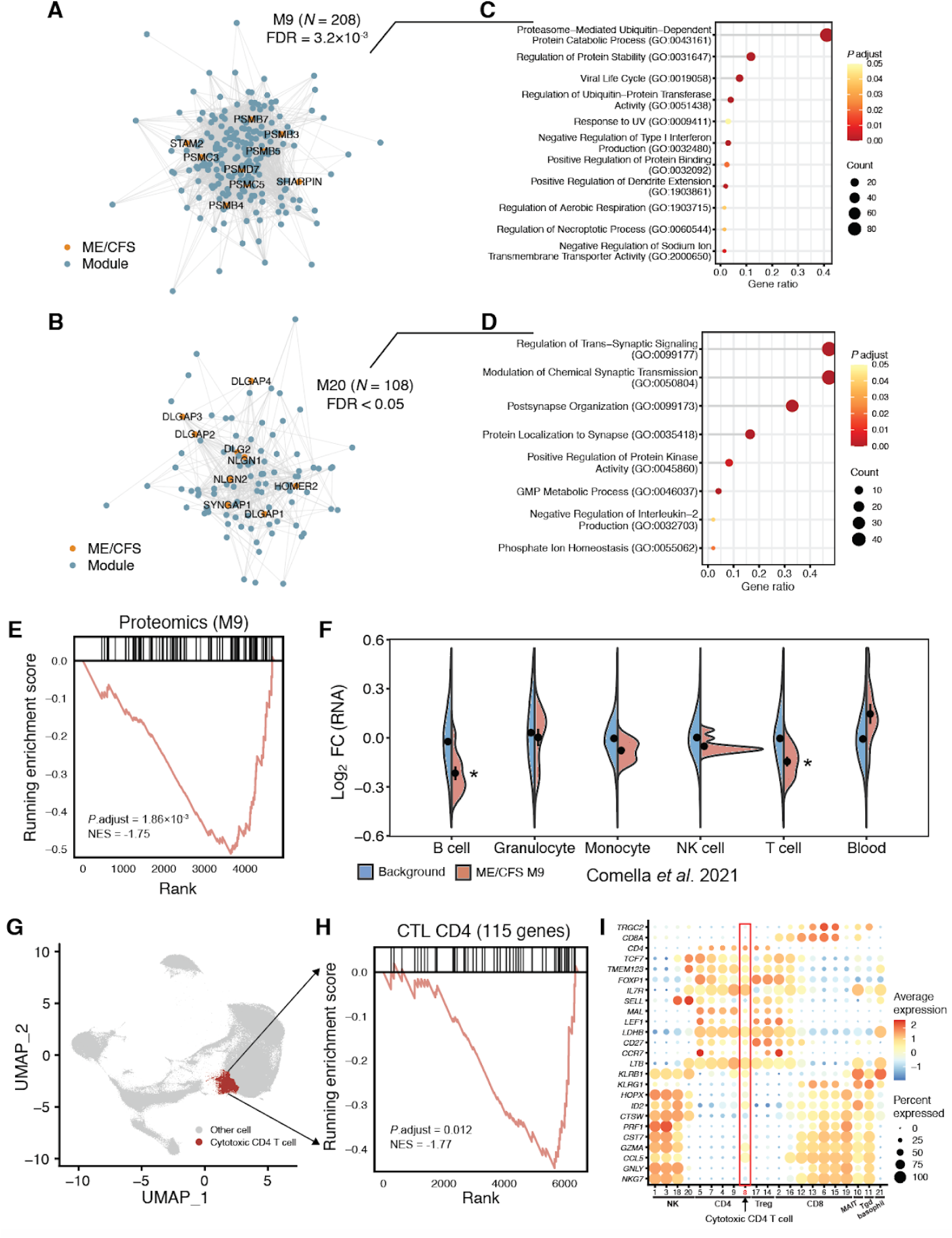
Network dissection and multi-omic analysis of ME/CFS genes. (A-B) Gene modules, including M9 (A) and M20 (B), enriched with ME/CFS genes. FDR, false discovery rate. *P*-value by Fisher’s exact test. (C-D) Gene ontology (GO) analysis (biological process) for M9 (C) and M20 (D) genes. Redundant GO terms were removed using the “simplify” function provided by “clusterProfiler”^36^. GO terms with adjusted *P* < 0.05 were visualized. *P*-value by two-sided Fisher’s exact test. (E) GSEA running enrichment plot of plasma proteomics against M9 genes. Negative enrichment score indicates downregulation in ME/CFS and vice versa. GSEA, gene set enrichment analysis; NES, normalized enrichment score. (F) Gene expression comparison for M9 ME/CFS genes between patients and healthy individuals across multiple blood cell types. Dot and error bar represent mean and standard error, respectively. *P*-value by two-sided Wilcoxon rank-sum test followed by Bonferroni correction. FC, fold change; NK, natural killer; *, adjusted *P* < 0.05. (G) Uniform Manifold Approximation and Projection (UMAP) plot of scRNA-seq data, with the cytotoxic CD4 T cell cluster highlighted in red. (H) GSEA running enrichment plot of ME/CFS genes in scRNA-seq pseudobulk data from the cytotoxic CD4 T cell cluster. Negative enrichment score indicates downregulation in ME/CFS cases compared to healthy controls, and vice versa. CTL, cytotoxic T lymphocyte. (I) Relative expression of general, CD4-specific, and cytotoxic T cell marker genes (y-axis) per T cell cluster. Dots represent average expression (color) and percentage of expressing cells (size). Treg, regulatory T cell; mucosal-associated invariant T cell; Tgd, gamma delta T cell.

### ME/CFS genes are differentially expressed in multiple conditions

Having uncovered the important role of ME/CFS genes in normal tissues and cell types, we next aimed to assess their functional change in diseases, especially ME/CFS.

We first examined the protein expression of our ME/CFS genes based on a previously generated plasma proteome dataset^31^. Of the 115 genes, 57 corresponding proteins were measured in plasma of 20 ME/CFS subjects and 20 controls. We used gene set enrichment analysis (GSEA), which assesses if gene sets are significantly altered as a group, to determine if the ME/CFS genes or the significant modules were also dysregulated in ME/CFS patients compared to controls at the protein level. We found that the protein levels in module M9 (**Fig. 4A**) were significantly decreased in ME/CFS patients versus controls (adjusted *P* < 0.002, normalized enrichment score (NES) = –1.75; **Fig. 4E**). Of the nine ME/CFS genes in M9, four proteins were measured. Two of these, PSMB4 and PSMB5 (components of the 20S core proteasome complex), were part of the leading edge subset (i.e., the proteins that contributed the most to the enrichment signal). The 115 ME/CFS genes and other modules did not show significant enrichment (**Supplementary Fig. 4**).

We also investigated the expression patterns of ME/CFS genes across multiple blood cell types within ME/CFS patients using an RNA-seq dataset^32^ generated by Comella et al. Although no difference in expression of overall ME/CFS genes was observed (**Supplementary Fig. 5A**), ME/CFS genes within module M9 (**Fig. 4A**) were markedly down-regulated specifically in patient B cells and T cells compared to those from healthy samples (adjusted *P* < 0.05, two-sided Wilcoxon rank-sum test followed by Bonferroni correction; **Fig. 4F**; **Methods**), showing concordance with the protein dataset and underscoring their cell-type-specific disease relevance. As a negative control, no other module ME/CFS genes were differentially expressed within any patient cell types (*P* > 0.05, two-sided Wilcoxon rank-sum test; **Supplementary Fig. 5B-D**).

To explore the functional changes of the 115 ME/CFS genes with increased granularity, we performed GSEA in cell subsets generated with high resolution clustering of T lymphoid cells from single-cell RNA sequencing (scRNA-seq) of PBMCs derived from ME/CFS patients and controls^33,34^ (**Supplementary Fig. 6**). Starting with data from the T cell subset pseudobulked by sample and cell cluster, we again compared expression levels in ME/CFS versus controls for the 115 genes and significant modules. Due to the sparsity of scRNA-seq data, we filtered the genes included in the GSEA to the top quartile to avoid comparing low count genes between groups. Strikingly, the set of 115 ME/CFS genes were significantly downregulated in the cytotoxic CD4 T cell cluster (adjusted *P* < 0.02, NES = –1.77; 61/115 genes were included in GSEA; **Fig. 4G-H)**, identified as such due to its substantially higher expression of effector genes like *CCL5* and *GZMA* compared to conventional CD4 T cell subsets^35^ (**Fig. 4I**). 18 of these genes were found in the core enrichment driving the downregulation in ME/CFS patients (**Supplementary Table 3**). Although we did not find significant downregulation of M9 in any of these T lymphoid cell clusters, *SHARPIN* and *PSMD7*, both members of M9, were among the highest ranked genes driving enrichment. Other notable leading edge genes contributing to downregulation in ME/CFS patients include *DNMT3A*, which had the highest attention difference (case vs. controls) in the HEAL2 model, and *PTPN11,* which was ranked third.

To obtain insights into the functional role of ME/CFS genes in other diseases, we systematically analyzed blood RNA-seq data for 59 diseases^22^. Notably, the genes we associated with ME/CFS genes tended to be down-regulated in immunological and infectious diseases (*P* < 0.05, two-sided *t*-test; **Supplementary Fig. 7**; **Methods**), including streptococcal soft tissue infection, pneumococcal pneumonia, pediatric systemic inflammatory disease, and staphylococcus aureus bacteremia. This result, along with the negative correlation between HEAL2 risk score and fewer viral infections (**Fig. 1D**), implies linkage between infection and ME/CFS^6^.

These data together reinforce the implication of LoF in ME/CFS risk genes as potential upstream risk factors for ME/CFS.

### Genetic correlation between ME/CFS and other diseases

Patients with ME/CFS often exhibit symptoms that overlap with other conditions, and co-morbidities are commonly observed. This led us to question whether these co-morbidities contribute to the underlying disease mechanism or are downstream effects of pathogenesis. To investigate this, we systematically mapped the genetic correlations between ME/CFS and other complex or Mendelian disorders.

First, leveraging rare variant association studies^37^ for 4,529 diseases and traits in the UK Biobank (UKBB), we assessed the distribution of SKAT-O *P*-values of our ME/CFS genes per disease or trait (**Methods**), defining a genetic correlation if this distribution was significantly shifted from the background. Notably, we found that ME/CFS was genetically correlated with various complex diseases and traits in rare variants (*P* < 0,05, one-sided Wilcoxon rank-sum test; **Fig. 5A**), including depression, irritable bowel syndrome (IBS), and COVID-19 susceptibility (C2). Similar results were obtained based on the burden tests^37^ (**Fig. 5B**). Our results provide a rare-variant-based genetic linkage between ME/CFS and depression.

**Figure 5.**
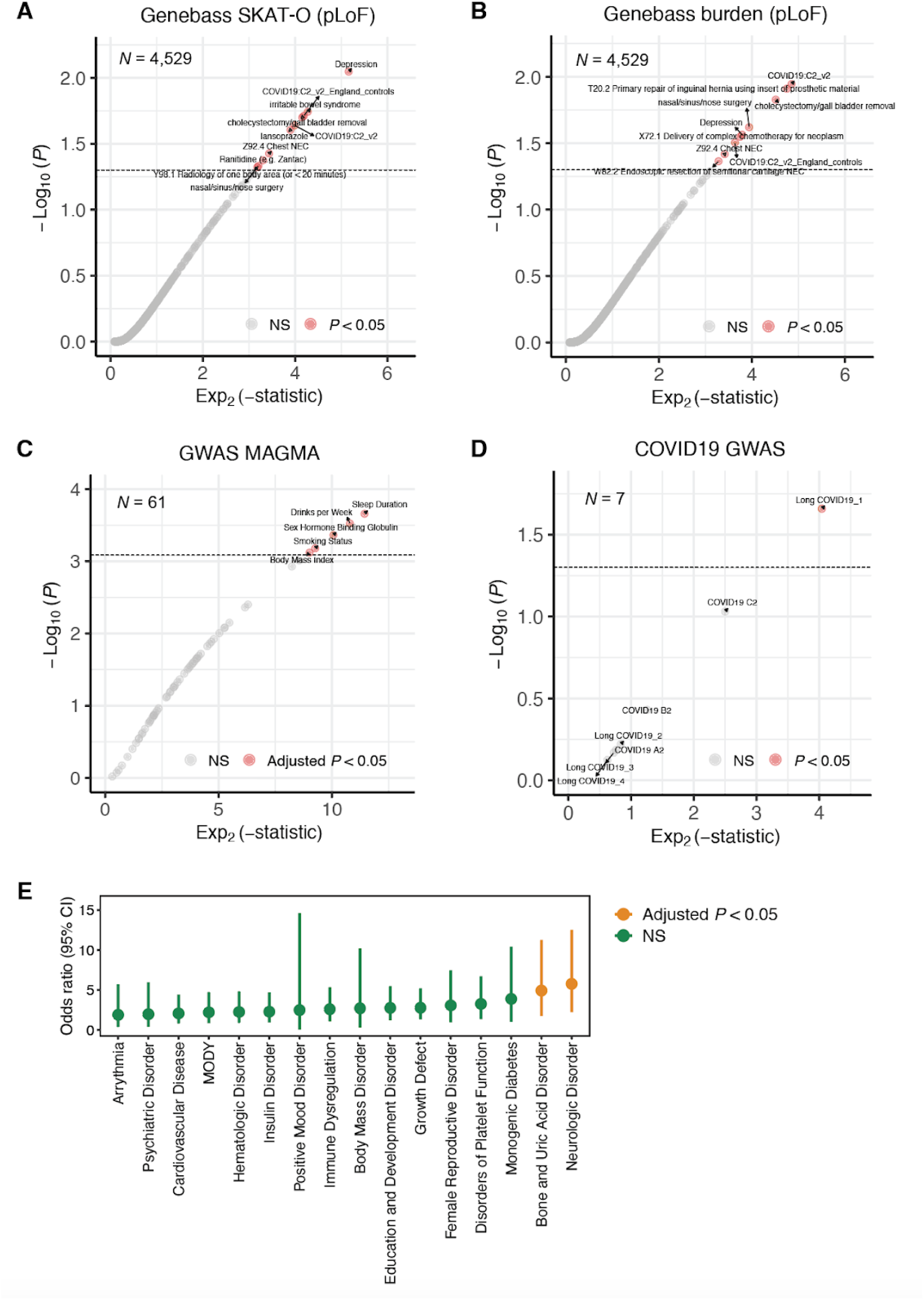
Genetic correlation between ME/CFS and other diseases and traits. (A-B) Rare-variant-based genetic correlation based on SKAT-O (A) and burden (B) tests. *P*-value by one-sided Wilcoxon rank-sum. pLoF, predicted loss-of-function; NS, not significant. (C-D) Common-variant-based genetic correlation based on GWAS for complex diseases and traits (C) and COVID-19 phenotypes (D). Bonferroni procedure was used for adjusting *P*-values. COVID19 A2, B2, and C2 indicates severe covid vs. population, hospitalized covid vs. population, and covid vs. population^42^, respectively; long COVID19_1, long COVID19_2, long COVID19_3, and long COVID19_4 indicate strict case vs. broad control, broad case vs. broad control, strict case vs. strict control, and broad case vs. strict control^38^, respectively. (E) Genetic correlation between ME/CFS and Mendelian disorders. *P*-value by Fisher’s exact test followed by Bonferroni correction. NS, not significant; CI, confidence interval; MODY, maturity-onset diabetes of the young. The dot and error bar indicate the odds ratio and 95% CI, respectively.

Next, we explored the common-variant-based genetic correlations based on the genome-wide association studies (GWASs) on 61 complex diseases and traits using a similar procedure (**Methods**). Interestingly, ME/CFS exhibited the strongest genetic correlation with sleep duration (adjusted *P* < 0.05, one-sided Wilcoxon rank-sum test followed by Bonferroni correction; **Fig. 5C**). When linked to COVID-19 phenotypes, we observed a significant common-variant-based genetic correlation between ME/CFS and long COVID-19^38^ (strict case definition; *P* < 0.05, one-sided Wilcoxon rank-sum test; **Fig. 5D**; **Methods**). This result is consistent with the symptom similarities between long COVID-19 and ME/CFS^39,40^, but provides a genetic perspective.

Finally, we investigated the genetic overlap between ME/CFS and 16 Mendelian disorders. In particular, we estimated the enrichment of ME/CFS genes within different Mendelian disease gene sets^41^ (**Methods**), where two disorders including neurologic disorder, and bone and uric acid disorder displayed significant overlaps with ME/CFS (adjusted *P* < 0.05, two-sided Fisher’s exact test followed by Bonferroni correction; **Fig. 5E**).

### ME/CFS genes reveal diverse disease-relevant mechanisms

Multiple genes prioritized by HEAL2, including *ACE*, *NAMPT*, and *IL12A*, are of biological and therapeutic relevance to ME/CFS pathology. *ACE* plays a critical role in vascular function and immune modulation, with any dysregulation impacting blood flow, inflammation and autonomic function, all of these are implicated in driving ME/CFS pathophysiology^43^. *ACE* has been linked to long COVID-19 and other post-viral syndromes^44^. Furthermore, the availability of well-established ACE modulators^45^ could alleviate vascular and inflammatory symptoms, making it an actionable target to pursue. *NAMPT*, contributing to significant downregulation of the 115 ME/CFS genes in cytotoxic CD4 T cells (**Fig. 4G**), is well-established as pivotal in NAD+ biosynthesis and linked to fatigue and mitochondrial dysfunction^46^. This role of *NAMPT*, linking mitochondrial function, energy metabolism, and immune regulation, indicates that treatments involving NAD metabolism warrant further investigation^47^. IL12A is a key cytokine in pro-inflammatory and Th1 immune responses, with dysregulation tied to immune abnormalities in ME/CFS and long COVID-19^48^..

Additionally, we also emphasize the relevance of *PSMB4, PSMB5*, *PSMB7*, *PSMD7, DNMT3A*, *NME1*, *NRAS* and *SYNGAP1* to the pathophysiology of ME/CFS. PSMB4, PSMB5, PSMB7, and PSMD7, all proteasome subunits, are integral to antigen processing and immune regulation, making them highly relevant to ME/CFS’s immune dysfunction and persistent inflammation^49^. In our GSEA analyses, PSMB4 and PSMB5 proteins were driving downregulation of functional module M9 (**Fig. 4A**) in plasma of ME/CFS patients versus controls, while *PSMD7* contributed to downregulation of the 115 ME/CFS genes in cytotoxic CD4 T cells (**Fig. 4E** and **4G**). Targeting the proteasome complex using proteasome modulators offers potential for modulating immune responses and alleviating systemic inflammation^50^. *NME1* is directly involved in mitochondrial function and systemic energy production^51^. *NRAS*, via RAS/MAPK signaling, ties into immune dysregulation and chronic inflammation, presenting a potential pathway for anti-inflammatory interventions^52^.

As highlighted in our network analysis, ME/CFS genes participate in biological pathways associated with synaptic function (M20; **Fig. 4B** and **4D**). *SYNGAP1,* another gene driving downregulation of the ME/CFS genes in cytotoxic CD4 T cells (**Fig. 4G**) and a member of M20, is involved in synaptic signaling and plasticity, essential for brain function with mutations linked to neurodevelopmental and psychiatric disorders^53^. *SYNGAP1*’s role in synaptic signaling highlights its potential connection to neurological symptoms in ME/CFS, offering therapeutic

potential in neuroprotective strategies^54^. Neuroligins NLGN1 and NLGN2^55^, DLG2^56^, GRM1^57^, and the DLGAP family including DLGAP1, DLGAP2, DLGAP3, and DLGAP4^58^ are proteins that regulate synaptic function and glutaminergic synaptic function in particular. Both the neuroligins and the DLGAP proteins are postsynaptic proteins with a role in synapse organisation^59^. Neuroligins bind to presynaptic neurexins and have a role in synapse specification. This entire set of synaptic genes has been linked to various neuropsychiatric disorders including clinical depression^59^. *NRGN1* was the only genome-wide significant hit in a GWAS of suicide behaviour that is linked to major depression^60^. A truncating LoF mutation within *NRGN1* has been linked to depression in the context of familial Alzheimer’s disease^61^. Most prominently, this set of synaptic genes is implicated in attention deficit hyperactivity disorder (ADHD) and autistic spectrum disorder (ASD) (Wikipathways WP5420). Indeed the entire set of ME/CFS genes is significantly enriched in this pathway (*P* = 3.9 x 10^-10^, two-sided Fisher’s exact test). A shared genetic basis between ME/CFS and ADHD/ASD suggests that these two disorders may have shared aspects of molecular dysfunction. It is intriguing that the typical age of onset of ADHD/ASD is in childhood whereas ME/CFS typically manifests in adolescence or adulthood^62^. If both disorders share a related mechanism then perhaps they are age-specific manifestations of a similar underlying process.

Collectively, this expanded gene set integrates immune, metabolic, and neurological mechanisms, providing a multifaceted framework for understanding ME/CFS pathology and potential for developing targeted therapeutic interventions.

## Discussion

Our study represents a significant advancement in the understanding of the genetic underpinnings of ME/CFS. We conducted a large whole-genome sequencing analysis of ME/CFS by applying a novel deep learning framework, HEAL2. By leveraging whole genomes from multiple cohorts for rare variant analysis, we identified 115 risk genes associated with ME/CFS, many of which exhibit significant intolerance to loss-of-function mutations, which is corroborated using evidence from patient multi-omics analysis. These risk genes play critical roles in diverse tissues, including the central nervous system and immune system, aligning with the multisystemic nature of the disease^63^. Collectively, our study provides novel insights into the genetic etiology of ME/CFS and also highlights the power of integrating genome-wide data with machine learning approaches for rare variant analysis.

Our findings build upon earlier studies suggesting a genetic susceptibility in ME/CFS, and substantially extend this knowledge from prior studies, which were often constrained by small sample sizes and inconsistent methodologies. By employing whole-genome sequencing and integrating advanced computational methods like HEAL2, we have overcome many limitations of earlier research. HEAL2 identified 115 risk genes that exhibit intolerance to loss-of-function mutations and emphasizes the potential role of these genes in maintaining cellular and systemic homeostasis. This intolerance to loss-of-function mutations provides information about underlying disease mechanisms contributing to the pathogenesis of ME/CFS and experimentally validating these insights will open avenues for targeted therapeutic interventions aimed at modulating these pathways.

Network analyses further revealed these ME/CFS risk genes’ roles in diverse cellular processes and implicated multiple tissues in disease pathogenesis. Functional analysis uncovered significant expression of ME/CFS genes in critical tissues, such as the central nervous system (CNS), skeletal muscle, and immune cells. Overall, these findings align with the clinical manifestations of ME/CFS, including cognitive impairment, fatigue, and immune dysfunction.

The downregulation of ME/CFS genes and the differential expression patterns observed from patient multi-omics data support their functional relevance to disease mechanisms and their potential to be identified as potential therapeutic targets. Moreover, based on HEAL2-identified ME/CFS genes we uncovered genetic correlations between ME/CFS and conditions such as depression and long COVID-19 susceptibility. This result aligns with a recent GWAS^64^ linking long COVID-19 to depression and ME/CFS. These correlations offer a genetic rationale for observed clinical overlaps and suggest shared pathophysiological mechanisms, potentially rooted in immune and neuroinflammatory pathways.

HEAL2 extends our previous work of HEAL^12^, which is a rare-variant-based genetic risk predictor. Different from HEAL, HEAL2 extensively explores PPIs which could contribute to disease risk in addition to linear genotype-phenotype (G2P) relationships. We demonstrated, in the context of ME/CFS, HEAL2 significantly outperformed HEAL in disease prediction, which implies that the nonlinear gene interactions may shape a considerable genetic architecture of ME/CFS. In our recent work^65^, we also proved that the incorporation of PPIs could improve genetic risk prediction based on common variants.

Given the well-established interpretation of the coding variants, in this study we only focused on missense and LoF variants in the analysis. However, noncoding variants, which account for a large proportion of human genetic variants, are missing in our current modeling. It is still a challenge in the field^66^, compared to coding variants, to interpret the function of noncoding genomes and variants, given their functional heterogeneity and dynamics across different tissues and conditions^67^. On the other hand, HEAL2 can be easily extended to incorporate noncoding variants in disease prediction, if the noncoding variant effects and their target genes are well defined. We anticipate that, along with the progress in the study of noncoding variants, HEAL2 could be applied to model G2P more comprehensively by integrating both common and rare, coding and noncoding variants.

Although gene discovery power has been enhanced by our machine learning modeling, the generalizability and stability of genetic risk prediction need further validation on additional independent cohorts. In this study, we only focused on European samples using a stringent cutoff, aiming to remove latent population-related genetic factors that could bias our biological discovery. However, it is well known that the genetic architecture of complex diseases and traits diverge among populations^68^. Thus it remains an open question whether the same conclusions hold for ME/CFS patients from other diverse populations. The solution of this question requires future work that focuses on including non-EUR ME/CFS cohorts.

This study highlights the power of integrating machine learning with genomics for understanding complex diseases. HEAL2 not only identifies genetic factors contributing to ME/CFS but also provides a framework for rare variant analysis applicable to other complex heterogeneous conditions. The identification of specific genetic variants and pathways involved in ME/CFS pathogenesis has important clinical implications for diagnostics and therapeutics. This knowledge could accelerate the discovery of biomarkers and therapeutic targets, moving toward precision medicine for ME/CFS and related disorders. The genetic correlations with other diseases suggest promising avenues for repositioning existing therapies or developing targeted treatments that address these overlapping mechanisms. More experimental follow-ups are needed to confirm our data-driven implications.

In conclusion, our study provides a genetic characterization of ME/CFS and reveals novel insights into its pathophysiology. The HEAL2 framework offers a robust analytical framework for rare variant analysis, promising to enhance our understanding of complex diseases.

## Data Availability

The whole-genome sequencing data have been made available on Synapse (Project SynID: syn64610522). All other data used in this study are publicly available and can be accessed through the original publications.

## Code Availability

The source code of HEAL2 has been uploaded in the Supplementary Materials.

## Acknowledgements

We are sincerely grateful for all research participants, including ME/CFS patients and healthy individuals, for their involvement and dedication. This study was carried out using samples from the CureME and UK ME/CFS Biobank (REC reference 21/WA/0388 to M.P.S.), which were processed and stored at the UCL-RFH BioBank. This study is supported by the Candace & Bert Forbes gift funds (to M.P.S.). M.J.Z. acknowledges the support of the Shurl and Kay Curci Foundation.

We thank the following individuals who participated in one or more of these activities to acquire the Cornell samples – participant screening, participant assessment, blood collection, data curation: Victoria Birdsall, John Chia, Patricia Doty, Carl Franconi, Tiffany Ong, Betsy Keller, Susan Levine, Xiangling Mao, Geoffrey Moore, Maria Russell, Dikoma Shungu, Jared Stevens, Kristin Treat, and David Wang.

NIH U54NS105541, an initial grant provided to the Cornell ME/CFS Collaborative Research Center, was co-funded by the National Institute of Neurological Disorders and Stroke, National Institute of Allergy and Infectious Diseases (NIAID), National Institute on Drug Abuse, National Heart, Lung, and Blood Institute, National Human Genome Research Institute, and the Office of the Director. A renewal grant was made to the Cornell Center from NIH NIAID: U54AI178855.

## Author Contributions

S.Z. and M.P.S. conceived and designed the project. S.Z., and M.K. designed HEAL2. M.K. implemented HEAL and HEAL2 with assistance from H.L. Sample preparation and processing was conducted by T.C. V.C. carried out data preprocessing, quality controls, and ancestry analysis. F.J. provided part of the Stanford samples. M.R.H. and A.G. provided the Cornell samples. M.L. and M.J.Z. performed MAGMA analysis. S.Z., M.K., V.C., K.A.G. and D.S.I. performed data analysis with the help from all other authors. All authors are responsible for data interpretation. M.P.S. and S.Z. supervised the project. S.Z., V.C., M.K., and K.A.G. prepared the manuscript with the help from all other authors.

## Competing Interests

M.P.S is a cofounder and scientific advisor of Crosshair Therapeutics, Exposomics, Filtricine, Fodsel, iollo, InVu Health, January AI, Marble Therapeutics, Mirvie, Next Thought AI, Orange Street Ventures, Personalis, Protos Biologics, Qbio, RTHM, SensOmics. M.P.S. is a scientific advisor of Abbratech, Applied Cognition, Enovone, Jupiter Therapeutics, M3 Helium, Mitrix, Neuvivo, Onza, Sigil Biosciences, TranscribeGlass, WndrHLTH, Yuvan Research. M.P.S. is a cofounder of NiMo Therapeutics. M.P.S. is an investor and scientific advisor of R42 and Swaza.

M.P.S. is an investor in Repair Biotechnologies.

M.R.H. is a member of the scientific advisory boards of the Open Medicine Foundation, Solve CFS/ME, the WE&ME Foundation, and Simmaron Research.

## Supplementary Figures

**Supplementary Figure 1. Simulation results.** (A-B) Prediction performance of five-fold cross-validation (100 repeats), evaluated using the receiver operating characteristic (ROC) curve (A) and the precision-recall curve (B). The curve and shaded area represent the AUROC mean and 95% confidence interval (CI), respectively. (C-D) Prediction performance of testing (500 repeats) on an independent dataset, evaluated using the ROC curve (C) and the precision-recall curve (D). (E-F) Performance of HEAL2 gene prioritization, evaluated using the ROC curve (E) and the precision-recall curve (F). The labels of 100 genes selected to generate the disease status in simulation were 1, and the rest of genes were labeled as 0.

**Supplementary Figure 2. Prediction performance evaluation in precision-recall curve.** (A) Prediction performance of five-fold cross-validation (100 repeats) on the discovery cohort. The curve and shaded area represent the AUPRC mean and 95% confidence interval (CI), respectively. AUPRC, area under the precision-recall curve; PC, principal component. (B) Prediction performance of testing (500 repeats) on the independent Cornell cohort. The curve and shaded area represent the AUPRC mean and 95% CI, respectively.

**Supplementary Figure 3. Network analysis for ME/CFS genes.** (A-B) Gene modules, including M15 (A) and M18 (B), enriched with ME/CFS genes. FDR, false discovery rate. *P*-value by Fisher’s exact test. (C-D) Gene ontology (GO) analysis (biological process) for M15 (C) and M18 (D) genes. Redundant GO terms were removed using the “simplify” function provided by “clusterProfiler”. GO terms with adjusted *P* < 0.05 were visualized. *P*-value by two-sided Fisher’s exact test.

**Supplementary Figure 4. Additional proteomics GSEA results for modules (A) M15, (B) M18, (C) M20, and (D) all 115 ME/CFS genes, respectively.** GSEA, gene set enrichment analysis; NES, normalized enrichment score.

**Supplementary Figure 5. Differential expression analysis of ME/CFS genes based on patient data.** (A-D) Expression comparison across multiple blood cell types between cases and controls for all ME/CFS genes (A), M15 ME/CFS genes (B), M18 ME/CFS genes (C), and M20 ME/CFS genes (D), respectively. Dot and error bar represent mean and standard error, respectively. No difference was significant by the two-sided Wilcoxon rank-sum test. NK, natural killer.

**Supplementary Figure 6. UMAP plot of the PBMC scRNA-seq dataset for ME/CFS.** Cells were clustered and annotated as described elsewhere^34^. UMAP, uniform manifold approximation and projection; PBMC, peripheral blood mononuclear cell; NK, natural killer cell; Th, T helper cell; CTL, cytotoxic T lymphocyte; Treg, regulatory T cell; MAIT, mucosal-associated invariant T cell. The figure was adapted from Iu et al.^34^

**Supplementary Figure 7. Gene expression patterns for ME/CFS genes in blood across 59 diseases.** NS, not significant. *P*-value by two-sided *t*-test.

## Supplementary Tables

**Supplementary Table 1. Prediction accuracy of different ME/CFS symptoms within the UK CureME cohort based on HEAL and HEAL2 risk scores. Acc, accuracy.**

**Supplementary Table 2. Genes prioritized by HEAL2.**

**Supplementary Table 3. 18 leading edge genes in GSEA of 115 ME/CFS genes in cytotoxic CD4 T cells.** GSEA, gene set enrichment analysis.

## Methods

### Stanford ME/CFS cohort

The Stanford ME/CFS cohort comprises 364 participants (208 cases and 156 controls), including members from 22 families totaling 74 participants, with 9 identical twins discordant for ME/CFS. Participants received ME/CFS diagnoses from specialized clinicians in the Bay Area and underwent comprehensive clinical assessments to confirm diagnoses based on ICC and IOM criteria^69–71^. Detailed phenotypic data were collected for all participants, ensuring a rich dataset for analysis. Blood samples were collected over several years at multiple Stanford locations and, for very severe and extremely severe cases, at patients’ homes. Samples were collected using 10 ml K2 EDTA vacutainers, processed immediately, snap-frozen in liquid nitrogen, and stored at –80°C. The research was approved by the Institutional Review Board at Stanford University (40146), adhering to rigorous protocols to ensure high-quality, anonymized data and samples.

We further increased the number of negative controls by incorporating samples from the iPOP (*N* = 110) and hPOP (*N* = 268) cohorts which are described elsewhere^72,73^. This yielded the final Stanford ME/CFS cohort including 208 patients and 534 controls.

### UK CureME ME/CFS cohort

The UK CureME cohort^74^ is a comprehensive resource designed to advance research into ME/CFS and is part of the UK ME/CFS Biobank initiative (https://cureme.lshtm.ac.uk/). Participants undergo thorough clinical assessments to confirm diagnoses and provide detailed phenotypic data. Biological samples, primarily blood, are collected at multiple time points to facilitate longitudinal analyses. The cohort adheres to rigorous protocols to ensure high-quality, anonymized data and samples. From the UK CureME cohort, we received blood samples for 190 individuals with ME/CFS and 30 healthy controls who provided consent for WGS (RFL Biobank Ethical Reference number: NC2021.24). The cohort adheres to rigorous protocols to ensure high-quality, anonymized data and samples, which are stored at the UCL/RFH Biobank in London, UK.

### Cornell ME/CFS cohort

The Cornell cohort was composed of 66 individuals with ME/CFS and 47 healthy controls who consented to provide blood for WGS. Almost all were participants in the cohort described by Moore et al.^75^, though a few provided blood but did not participate in exercise testing. Subjects visited three different sites for blood collection: ID Med in Torrance, CA; Cornell University and Ithaca College in Ithaca, NY; and Weill Cornell Medicine in New York, NY. Following blood draw into K2 EDTA vacutainers, samples were provided to processing labs at each site. 1 ml whole blood aliquots along with demographic information were provided to Stanford for isolation of DNA. The research was approved by Institutional Review Boards at Cornell University (IRB# 0001855), Ithaca College (IRB# 1017-12D), or Weill Cornell Medicine (IRB# 1708018518).

### HEAL2 model specification

To investigate the genetic architecture of ME/CFS, we developed HEAL2, an advanced deep learning framework that integrates graph neural networks (GNNs) with an attention-readout mechanism coupled with a sparse autoencoder (SAE) for interpretability. HEAL2 extends our previous linear model, HEAL^12^, by incorporating multidimensional pathogenicity scores and leverages a protein-protein interaction (PPI) network to capture complex, non-additive genetic interactions. Similar to PRS-Net^65^, a GNN model but for polygenic risk scores, we first derive our PPI network from the STRING database^30^, as protein-protein interactions are good representations of relevant gene-gene associations, selecting only for high confidence (score > 700) links. In detail, we build the graph 𝓖 = (𝓥, 𝜠), where 𝓥 is the set of nodes (genes) while 𝜠 corresponds to the set of edges (𝓋_𝑖_, 𝓋_j_), representing the interaction between gene 𝑖 and 𝑗 as obtained from the STRING database. Next, every node is further extended with a self-loop (𝓋_𝑖_, 𝓋_j_), allowing each gene to retain its own information during message passing, preserving individual gene characteristics alongside interaction effects.

To construct the input features for HEAL2, we began by representing each individual’s genetic data through a comprehensive feature vector of the mutational burden across genes. For every gene in each sample, we aggregated the rare missense variants and their corresponding annotated scores by both summing, capturing the cumulative mutational burden for each gene, and taking the maximum pathogenicity score across all variants, highlighting the potential impact of the most deleterious variant. For LoF variants (i.e., start loss, stop gain, stop loss, and frameshift indels), which may lack predicted scores, we also included the gene’s probability of being loss-of-function intolerant (pLI) score from gnomAD v.4.1. This process resulted in individual-specific gene feature vectors that capture both the cumulative and peak effects of rare coding variants for each gene across 39 annotated scores. Formally, each gene 𝓋_𝑖_ ∈ 𝓥 is assigned an initial feature vector 𝐡_𝑖_ ∈ 𝓗, where 𝓗 ∈ ℝ^|𝓥|×78^ and |𝓥| is the total number of genes in 𝓖. Our input gene feature vectors are first passed through a multilayer perceptron (MLP) to obtain initial gene embeddings:

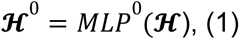

where 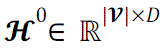 and D is the dimension of hidden states.

To capture nonlinear gene-gene interactions within our PPI network, we apply multiple layers of the Graph Isomorphism Network^76^ (GIN). These GIN layers execute message-passing operations across the network, progressively updating each node’s representation by incorporating information from its neighboring nodes. At each layer 𝑘, the representation of gene 𝓋_i_ is updated as follows:

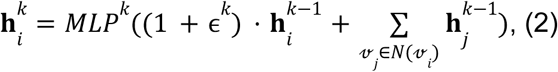

where 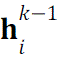 represents the hidden state of 𝓋*_i_* from the 𝑘 − 1-th layer, 𝑁(𝓋*_i_*) denotes the set of neighboring genes in the GGI network, and ɛ^𝑘^ is a learnable parameter specific to layer 𝑘. The 𝑀𝐿𝑃^𝑘^ transforms the combined information, allowing the model to weigh self-information and neighbor contributions at each layer adaptively. After 𝑘 iterations, each gene’s representation integrates information from genes within its 𝑘-hop neighborhood, embedding both local and extended gene interaction patterns.

For model interpretation and prioritizing genes contributing to ME/CFS risk, we employ an attentive readout module integrated with a sparse autoencoder (SAE). The attentive readout focuses on key genes by assigning attention weights, while the SAE refines these features by enforcing sparsity, highlighting the most informative genes. For each sample, the global representation 𝐡_𝑔𝑟𝑎𝑝ℎ_ is first derived through the attention mechanism as follows:

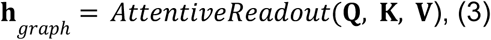

where 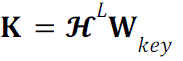 and 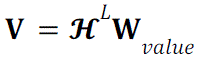 are the key and value matrices derived from the final GNN layer embeddings 𝓗^𝐿^ using trainable projection matrices 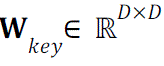 and 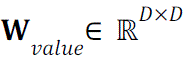.

The attention scores are calculated as:

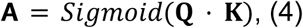

where 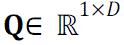 is a trainable query vector, assigning importance to each gene, with higher values in 𝝖 indicating greater relevance.

The attention-weighted hidden representations, 𝝖 · 𝐕, are subsequently passed through the SAE for further refinement. Within the SAE, the encoder maps these attention-weighted hidden features to a sparse latent space:

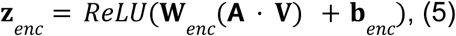

where 𝐖_𝑒𝑛𝑐_ and 𝐛_𝑒𝑛𝑐_ are the encoder’s weights and bias terms. To encourage sparsity, an 𝐿_1_ penalty is applied to 𝐳_𝑒𝑛𝑐_, helping to isolate the most informative genes. The latent representation is then decoded back to the original dimensionality and assigned 𝐡_𝑔𝑟𝑎𝑝ℎ_. The reconstruction error is combined with the sparsity constraint to form the following loss function:

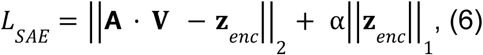

where α is a sparsity regularization coefficient and was set to 1 in this study.

Finally, the graph-level representation 𝐡_𝑔𝑟𝑎𝑝ℎ_, constructed from the SAE-processed embeddings across all nodes, is passed through an MLP, outputting the final ME/CFS classification. The training objective thus combines binary cross-entropy loss for classification and the SAE’s sparsity-constrained reconstruction loss, optimizing both prediction accuracy and interpretability. In this study, we used 78 input features to match our annotated scores while the rest of the model parameters were set to those of PRS-Net^65^ with 64 hidden dimensions, one gene-encoding layer, one GIN layer, and two predictor layers.

### HEAL model specification

To provide a fair comparison between HEAL2 and our previous model, HEAL^12^, we slightly extended it to include the multidimensional mutational burden used as input for HEAL2. Specifically, we redesigned HEAL to utilize the comprehensive gene feature vectors that capture both the cumulative and peak effects of rare variants across the 39 annotated scores for each gene. In our modification of HEAL, we employ a linear modeling approach that treats each gene independently, focusing on the aggregated mutational burden per sample without considering gene interactions. Formally, each gene 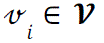 is assigned an initial feature vector 𝐡*_i_* ∈ ℝ^78^, identical to the input in HEAL2. Our model begins by transforming these gene-specific feature vectors into scalar gene scores using a linear transformation:

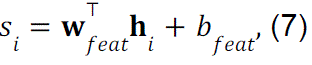

where 𝑠_𝑖_ ∈ ℝ is the score for each gene 𝓋*_𝑖_*, 𝐰_𝑓𝑒𝑎𝑡_ ∈ ℝ are the weights applied to the features, and 𝑏_𝑓𝑒𝑎𝑡_ is a bias term. This transformation effectively summarizes the multidimensional mutational burden of each gene into a single scalar value. Next, we aggregate the gene score into a single ME/CFS prediction score 𝑦 for each sample by applying another linear transformation:

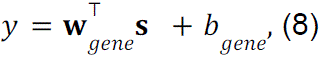

where 𝐬 ∈ ℝ^|𝓥|^ is the vector of gene scores, 𝐰*_𝑔𝑒𝑛𝑒_* ∈ ℝ are the weights assigned to each gene, and 𝑏*_𝑔𝑒𝑛𝑒_* is a bias term. As a key feature of the original HEAL model is applying LASSO regularization to force the model to focus on a key set of genes, we further introduce 𝐿_1_ regularization on both the feature-level weights 𝐰*_𝑓𝑒𝑎𝑡_* and the gene-level weights 𝐰*_𝑔𝑒𝑛𝑒_*, resulting in optimizing the final loss function:

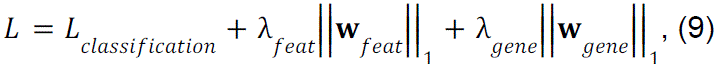

where 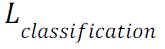 is the binary cross-entropy loss, while 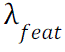 and 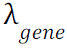 are hyperparameters controlling the strength of regularization at the feature and gene levels, respectively. After manual tuning on the validation set, we set 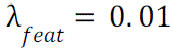 and 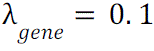.

### Model training and testing

We evaluated HEAL2’s ability to predict ME/CFS risk using both cross-validation within the discovery cohort (by combining the Stanford and UK CureME cohorts) and independent testing on the Cornell cohort. First, we applied stratified five-fold cross-validation (5-CV) on the discovery cohort, comprising 247 ME/CFS cases and 192 controls, to ensure that each fold retained the same proportion of cases and controls. This procedure, repeated 100 times with different random seeds, provides robust performance estimates by controlling for variance across different data splits.

The HEAL2 model was implemented in PyTorch with the Deep Graph Library (DGL). We trained the model using the AdamW optimizer, with a learning rate of 6.25 x 10^−5^, a batch size of 16 and weight decay of 1 x 10^−6^. To prevent overfitting, we employed early stopping, halting the training procedure if performance on the validation set failed to improve over 20 consecutive epochs.

For independent validation, we used the Cornell cohort (36 cases, 21 controls), withheld entirely from model training. Testing was performed when model training was complete on the discovery cohort, once every fold, resulting in 500 total tests on the independent cohort. Performance was evaluated by calculating the area under the receiver operating characteristic curve (AUROC) and the area under the precision-recall curve (AUPRC), with results reported as mean values and 95% confidence intervals across all repetitions.

Additionally, we evaluated the performance of our previous HEAL model using the same training and evaluation protocols for comparison. All experiments were conducted on multiple NVIDIA A100 GPUs.

### Simulation details

To validate HEAL2’s ability to model nonlinear genotype-phenotype relationships and accurately identify disease-relevant genes, we constructed a synthetic dataset with a “known” set of simulated causal genes. Specifically, we initialized HEAL2 with random, fixed weights and performed a forward pass of all samples to extract their final gene-level representations. From averaging these representations across all samples for each gene, we selected the top 100 genes to act as our ground-truth. We then amplified the influence of these 100 genes by scaling their specific weights in the final representation and repeated the forward pass. The resulting predictions from the model were thresholded at the median to assign simulated case/control status. The resulting synthetic dataset provided a controlled scenario with stable phenotypic labels related to a known set of 100 genes.

We then applied the same performance evaluation and gene prioritization procedures used in the ME/CFS analysis to this synthetic dataset for both HEAL2 and HEAL. To quantify how accurately HEAL2 recovered the artificially designated causal genes, we treated the known set of 100 “injected” causal genes as true positives and computed AUROC and AUPRC by comparing the attention-based gene prioritization against these ground-truth labels.

### DNA preparation and sequencing

Genomic DNA was isolated from whole blood using the QIAamp DNA Blood Midi Kit (Qiagen, Cat. No. 51185) following the manufacturer’s protocol. One milliliter of whole blood was used for each extraction. The isolated DNA was quantified using the Qubit™ dsDNA Quantification Assay Kit (Thermo Fisher Scientific, Cat. No. Q32851) and stored at –80°C until sequencing. Whole genome sequencing was performed by Novogene using their PCR-free WGS platform. For each sample, 300 ng to 1 μg of total genomic DNA was submitted. Prior to sequencing, all samples underwent rigorous quality control assessments to ensure only high-quality DNA was processed. Each sample was sequenced across 1-5 lanes to obtain more than 90 gigabases (Gb) pairs of reads per sample, and the resulting FASTQ files were concatenated. The sequencing data were deposited in an Amazon S3 bucket within the Stanford Data Ocean repository for secure storage and access. This comprehensive approach to DNA preparation and sequencing ensures high-quality data for downstream analyses, adhering to best practices in genomic research.

### Read alignment and variant calling

The read alignment and variant calling was done using the Sentieon’s DNAseq pipeline which adheres to GATK best practices while providing significant computational efficiency and accuracy. The pipeline implements the same mathematics used in the Broad Institute’s BWA-GATK HaplotypeCaller 4.0 workflow, but is highly parallelizable and optimized for performance, achieving 10X faster processing speeds for alignment. The pipeline processes data from FASTQ to VCF, completing BAM-to-VCF conversion in under 30 minutes. The pipeline supports extensive QC measures, such as metrics for base quality score distribution, read alignment quality, and duplicate marking, which were integrated seamlessly within the workflow.

### Quality controls

Following the best practice^12^, quality controls (QCs) for WGS were conducted using PLINK’s QC metrics^77^ which involves a series of steps to ensure that the data is clean, reliable, and suitable for downstream analysis. Sample-level and variant-level QCs were done where samples were filtered for high rate of genotype missingness, abnormal high or low heterozygosity rates or deviations from Hardy-Weinberg Equilibrium (HWE) to account for errors or poor-quality DNA. Closely related individuals were also filtered out by calculating Identity by Descent (IBD) and relatedness. Pruning was performed and individuals with first or second degree relative in the sample were removed.

### Ancestry analysis

Ancestry analysis was performed by leveraging the population structure information obtained from principal component analysis (PCA) using PLINK’s Eigensoft tool. PCA is a dimensionality reduction technique used in PLINK to summarize genetic variation into principal components (PCs). These PCs were plotted to visualize population structure, detect ancestry patterns, and spot any population outliers. We also performed population admixture analysis using the ADMIXTURE^78^ for ancestry inference and population structure analysis. The 1000 Genomes^79^ (1KG) were used as the reference to ADMIXTURE. Samples with EUR estimates *Q* > 0.99 were retained for downstream analysis.

### Variant annotation

Variant annotation was performed using Annovar^18^. Variant types, including missense, stop gain, stop loss, start loss, and frameshift indels, were annotated along with their affected genes. Allele frequencies were computed based on gnomAD^21^ v4.1. For pathogenicity scores for missense variants, we relied on the dbNSFP database^80^ which compiled numerous prediction scores.

### Input feature prioritization

HEAL2 evaluates the contribution of specific variant annotations and pathogenicity scores by calculating gradient-based feature importance. During model evaluation on the test set, the gradient of the model’s prediction with respect to the input features was computed for each sample. The absolute gradients were summed across all genes in the GGI network, condensing the overall contribution of each feature into a single importance score. For this analysis, we re-trained HEAL2 on the full cohort (Stanford, UK CureME, and Cornell) using a train-test-split of 80% for training and 20% for validation, repeated over 100 iterations employing the same early-stopping strategy as for the performance evaluation. Mean feature importance scores were computed per sample for each feature across all iterations to ensure stability. These scores were then compared between cases and controls, highlighting the features with the most relevance in regards to the classification.

### Gene prioritization

To further interpret HEAL2’s predictions, we prioritized genes contributing to ME/CFS risk using the model’s attentive readout mechanism. For each sample and gene, we took the absolute sum of the SAE-decoded attention-weighted representations across all hidden dimensions, generating a single score that condenses the gene’s contribution to ME/CFS classification. We then averaged these scores per sample across 100 iterations of training on the full cohort. The mean scores for each gene were compared between cases and controls using the Mann-Whitney U test, followed by Storey-Tibshirani adjustment^17^ for multiple hypothesis testing (*q*-value < 0.02). This process identified 115 genes with consistently higher attention scores in cases compared to controls, defining these as HEAL-2 prioritized ME/CFS risk genes.

### ME/CFS symptom analysis

To assess the relationship between genetic risk scores and ME/CFS symptoms, we analyzed HEAL2’s prediction scores in relation to additional phenotypic data from the UK CureME cohort. Specifically, we focused on the validation sets from the 100 iterations of 5-CV, calculating the mean prediction score for each sample across all iterations where the sample appeared in the validation set. Next, these averaged prediction scores were used to calculate the accuracy in capturing symptom-specific phenotypic variation for each binary phenotype. For comparison, this procedure was repeated using HEAL’s prediction scores to evaluate its performance on the same phenotypic data.

### LoF intolerance analysis

We used four different scores, including LoFtool^18^, RIVS^19^, pLI^20^, and LOEUF^21^, to quantify gene intolerance to LoF variants. Two-sided Wilcoxon rank-sum test was used to compare the scores of ME/CFS genes identified by HEAL2 with those of all background genes.

### Differential expression analysis

For differential expression (DE) analysis for baseline human tissues and cell types, we used a two-sided *t*-test to compare the expression levels of ME/CFS genes identified by HEAL2 with those of all expressed genes. The *t*-test statistics was used to estimate the expression difference, and the Bonferroni procedure was employed for multiple testing correction. Whenever there was no significance after *P*-value adjusting, we simply reported the raw *P*-values. Similarly, for DE analysis for disease data shown in Fig. 4F, we compared the fold changes of ME/CFS genes with those of all genes.

### Network analysis

We downloaded the human protein-protein interactions (PPIs) from STRING^30^ v12.0, comprising 19,622 proteins and 6,857,702 interactions. High-confidence PPIs (combined score >700) were extracted for downstream analysis, including 16,185 proteins and 236,000 interactions. To mitigate bias from hub proteins^81^, we applied the random walk with restart (RWR) algorithm with a restart probability of 0.5. This produced a smoothed network after retaining the top 5% predicted edges (*N* = 6,243,766). Next, we utilized the Louvain method^82^ to decompose the network into different modules. Following algorithm convergence, we obtained 1,261 modules with an average size of 13 nodes.

The enrichment of genes of interest within each module was tested using one-sided Fisher’s exact test. Modules with adjusted *P* < 0.05 based on Benjamini-Hochberg (BH) correction were considered significant.

### Gene ontology analysis

Gene ontology (GO) analysis was performed using Enrichr^83^ and clusterProfiler^36^.

### Gene expression enrichment analysis

Single cell RNA sequencing (scRNA-seq) of PBMCs from 30 ME/CFS patients and 30 healthy controls was performed as described in Vu et al.^33^ and Iu et al.^34^ Dimensional plots and marker plots of scRNA-seq were generated with Seurat v4.4.0. We obtained pseudobulk count data for this dataset from the authors for all samples and cell clusters generated in those publications. This included the initial clustering of all PBMCs into 29 clusters^33^ as well as 21 clusters from a high resolution re-clustering of T lymphoid cells^34^. Gene set enrichment analysis (GSEA) was performed comparing ME/CFS and controls within each gene cluster for custom gene lists including the 115 genes and the four significant modules from the network analysis (M9, M15, M18, M20). The input ranking is the Log2FC from DESeq2 analysis of the pseudobulk count data. The genes included in the GSEA ranked list for each cluster are in the top quartile based on mean normalized counts from DESeq2. GSEA was performed with the fgsea R package^84^ with the following parameters: eps = 0, minSize = 5, maxSize = 1000, nPermSimple = 10000. *P* values are determined by comparing each gene list to many permutations of random gene lists of the same size, followed by the Benjamini and Hochberg FDR correction procedure (*q* < 0.05).

### Proteomics enrichment analysis

Previously published^31^, fully normalized proteomics data was downloaded from mapmecfs.org. 4,790 unique human proteins were detected with the SomaScan aptamer-based assay in the plasma of 20 ME/CFS and 20 control subjects (all females). The 4,790 proteins included 57 of the 115 genes of interest identified here. GSEA was performed with the same gene lists and methods as above for the single-cell RNAseq data.The median Log2 fold change ME/CFS versus control was used as the input ranking for GSEA. For proteins with multiple measurements from different aptamers, the one with the highest |Log2FC| was used to prevent duplicates.

### MAGMA analysis

Gene-level analyses of GWAS data were conducted using MAGMA v1.10^85^. GWAS variants were mapped to genes using an annotation window of 10kb both upstream and downstream of the gene body. Linkage disequilibrium (LD) was adjusted based on the 1KG EUR panel. The Mean model (snp-wise=mean) was applied to derive the gene-level *P*-values.

### Genetic correlation analysis

For rare variant association study and GWAS data, we compared the *P*-values of ME/CFS genes with those of all background genes using a one-sided Wilcoxon rank-sum test. The Bonferroni procedure was adopted for *P*-value adjusting when available; otherwise the raw *P*-values were reported. For Mendelian disorder gene sets, we used a one-sided Fisher’s exact test to evaluate the enrichment of ME/CFS genes within each disease gene set, followed by the Bonferroni correction.

