## Supplementary figures and images for "Dissecting the genetic complexity of myalgic encephalomyelitis/chronic fatigue syndrome via deep learning-powered genome analysis"

### Supplementary Fig. 1

#### Supplementary Figure 1

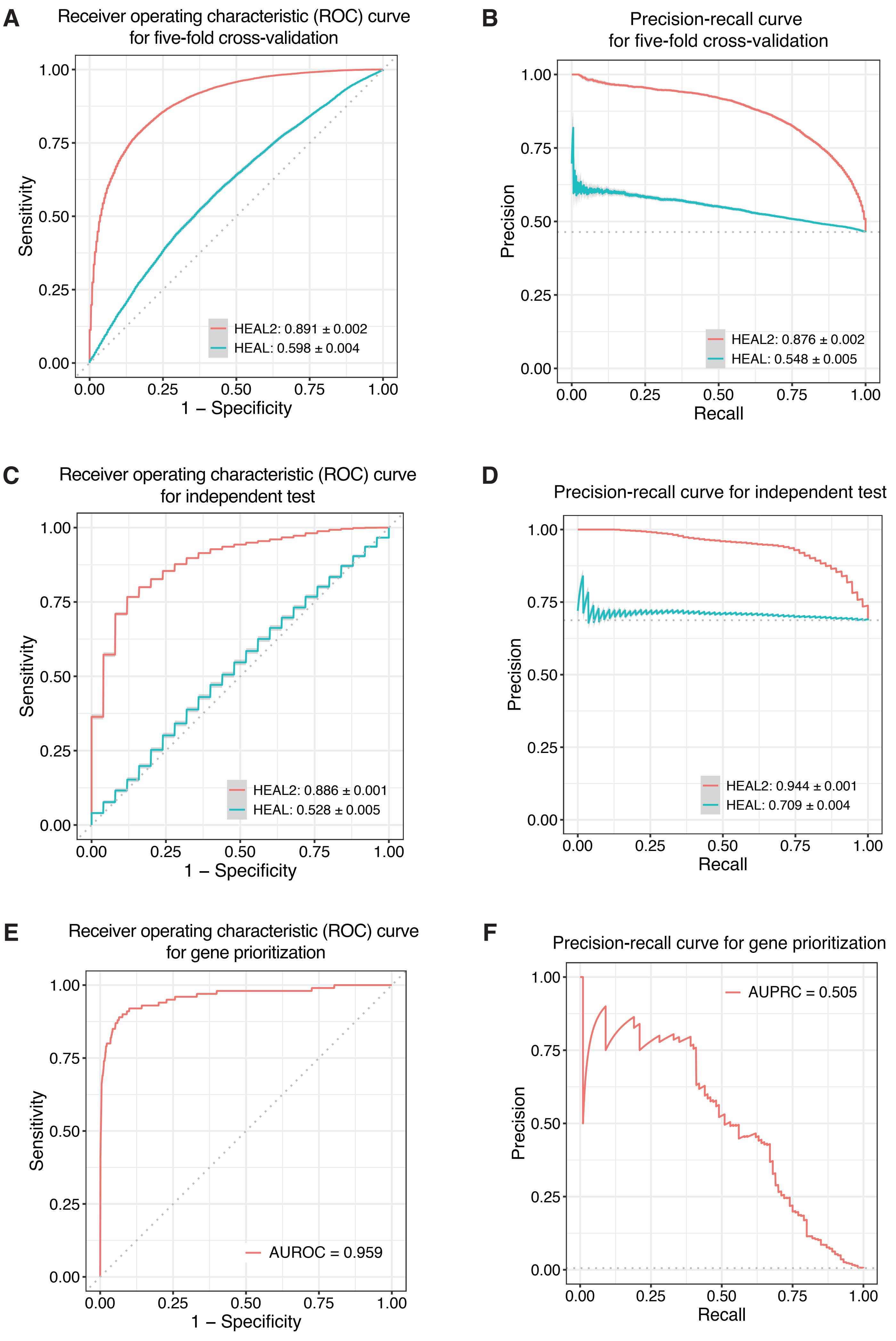

### Supplementary Fig. 2

##### Supplementary Figure 2

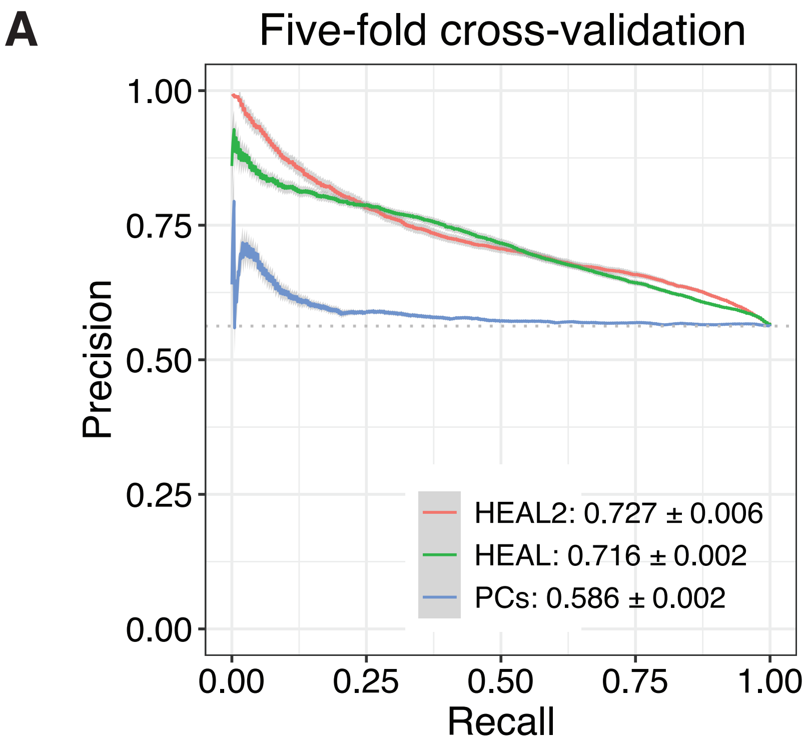


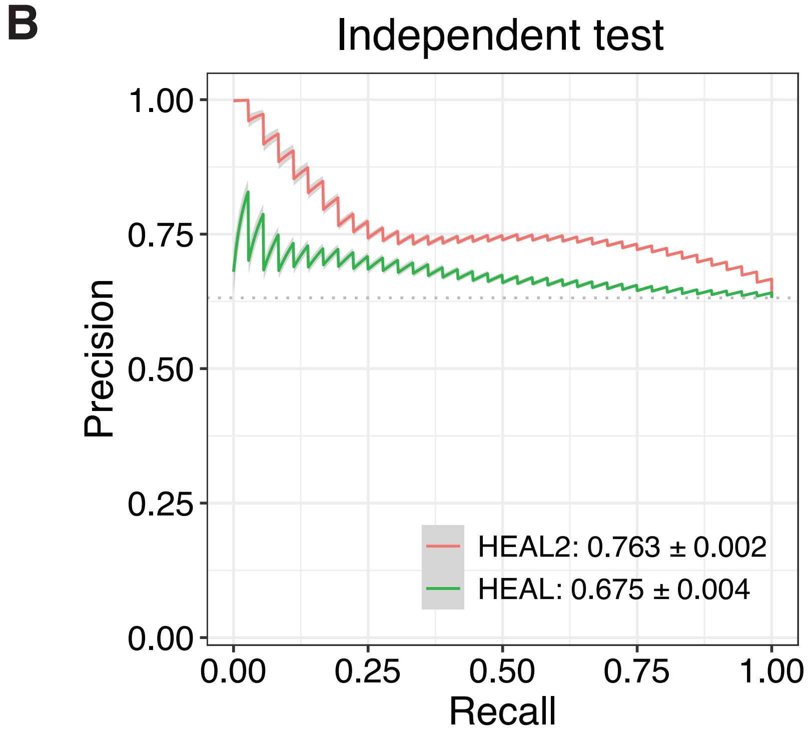

### Supplementary Fig. 3

#### Supplementary Figure 3

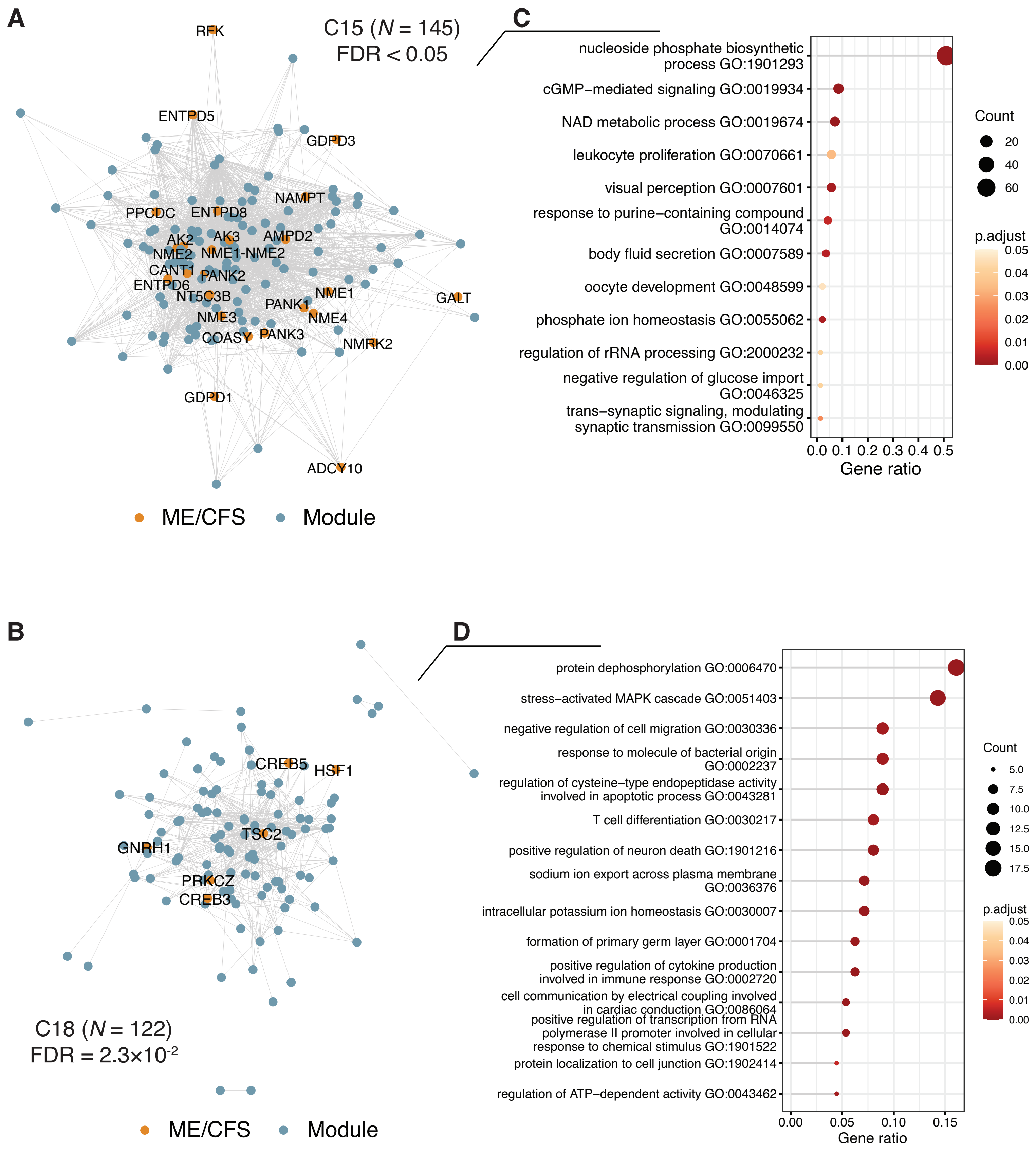

### Supplementary Fig. 4

##### Supplementary Figure 4

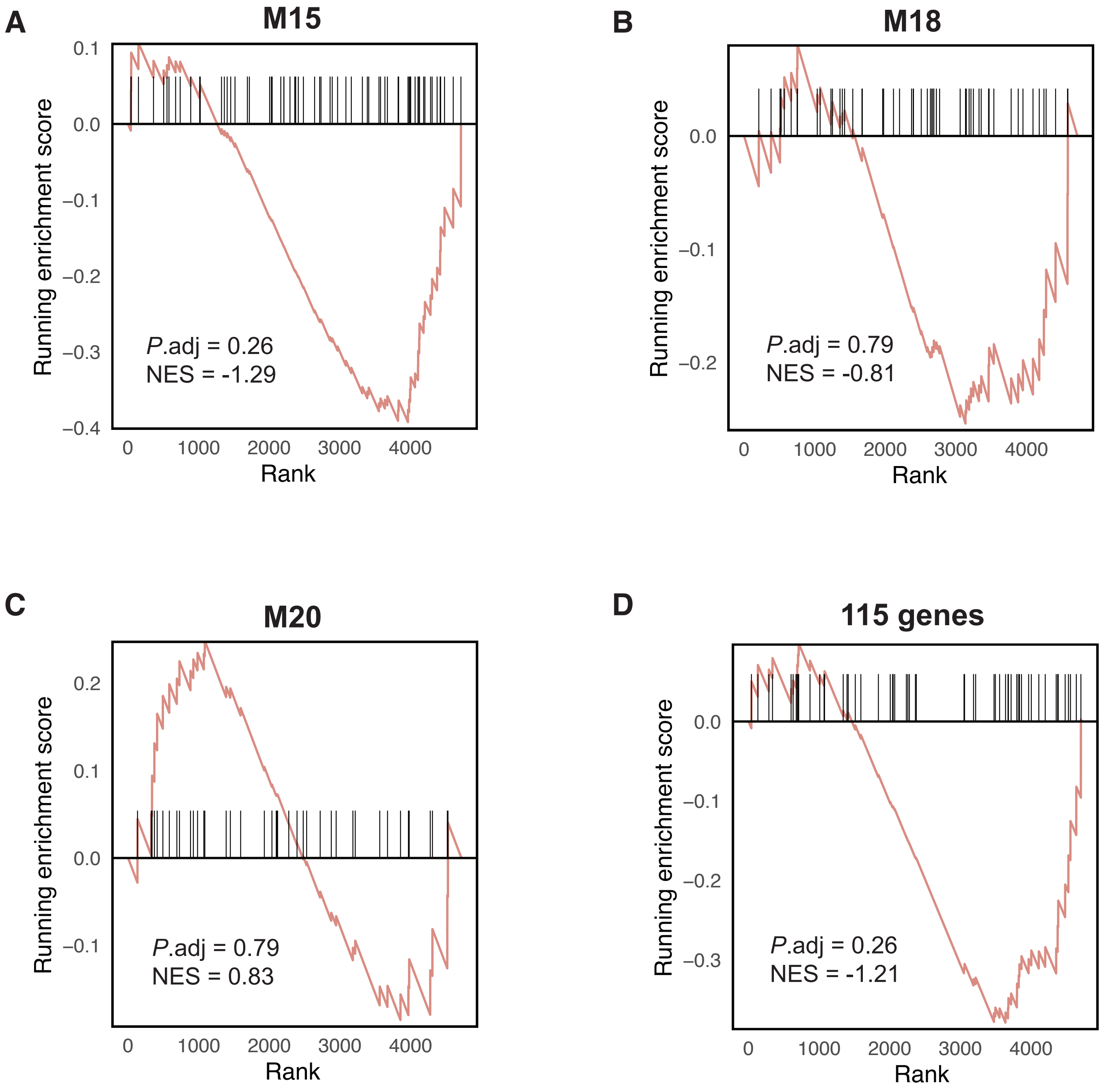

### Supplementary Fig. 5

#### Supplementary Figure 5

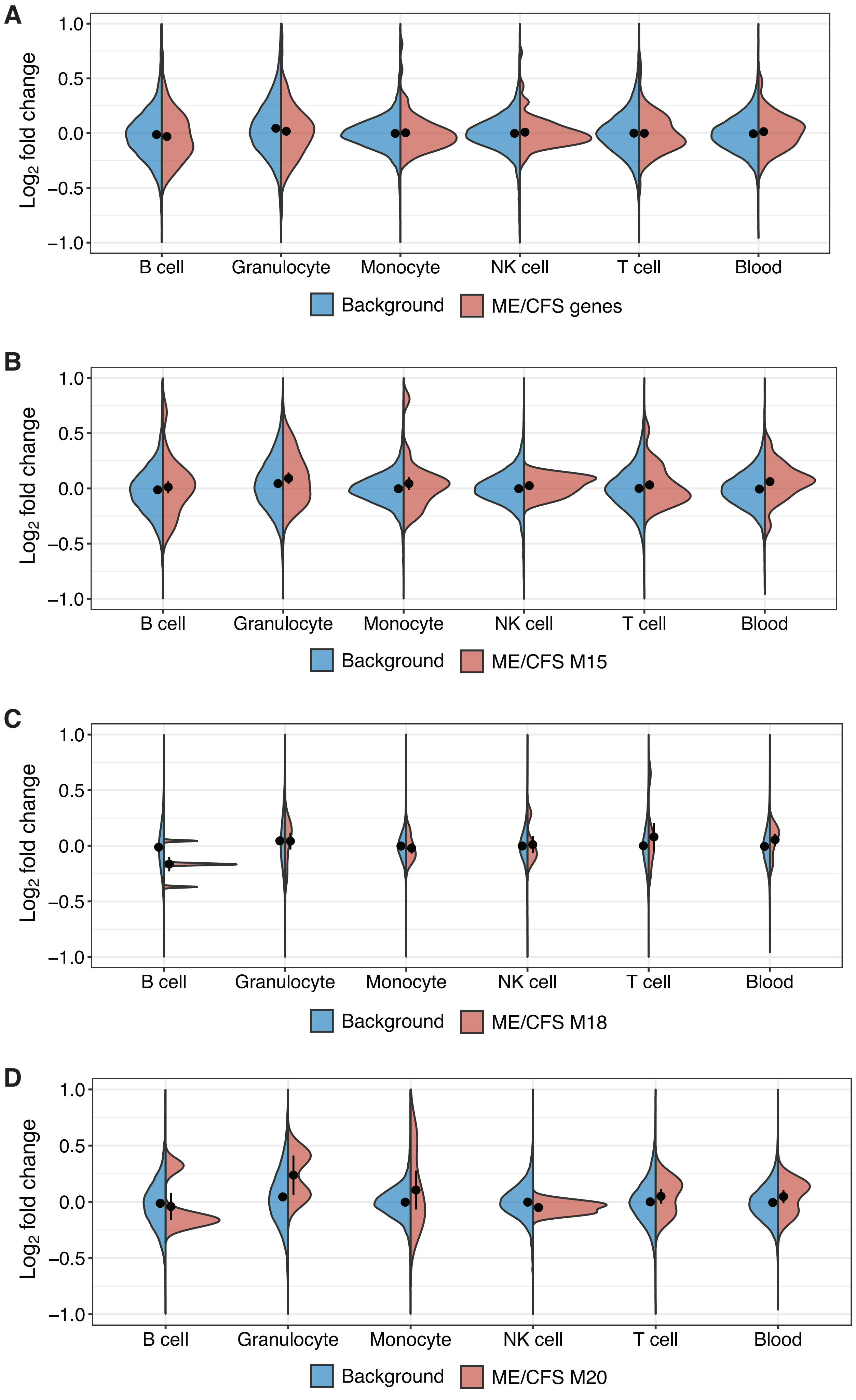

### Supplementary Fig. 6

#### Supplementary Figure 6

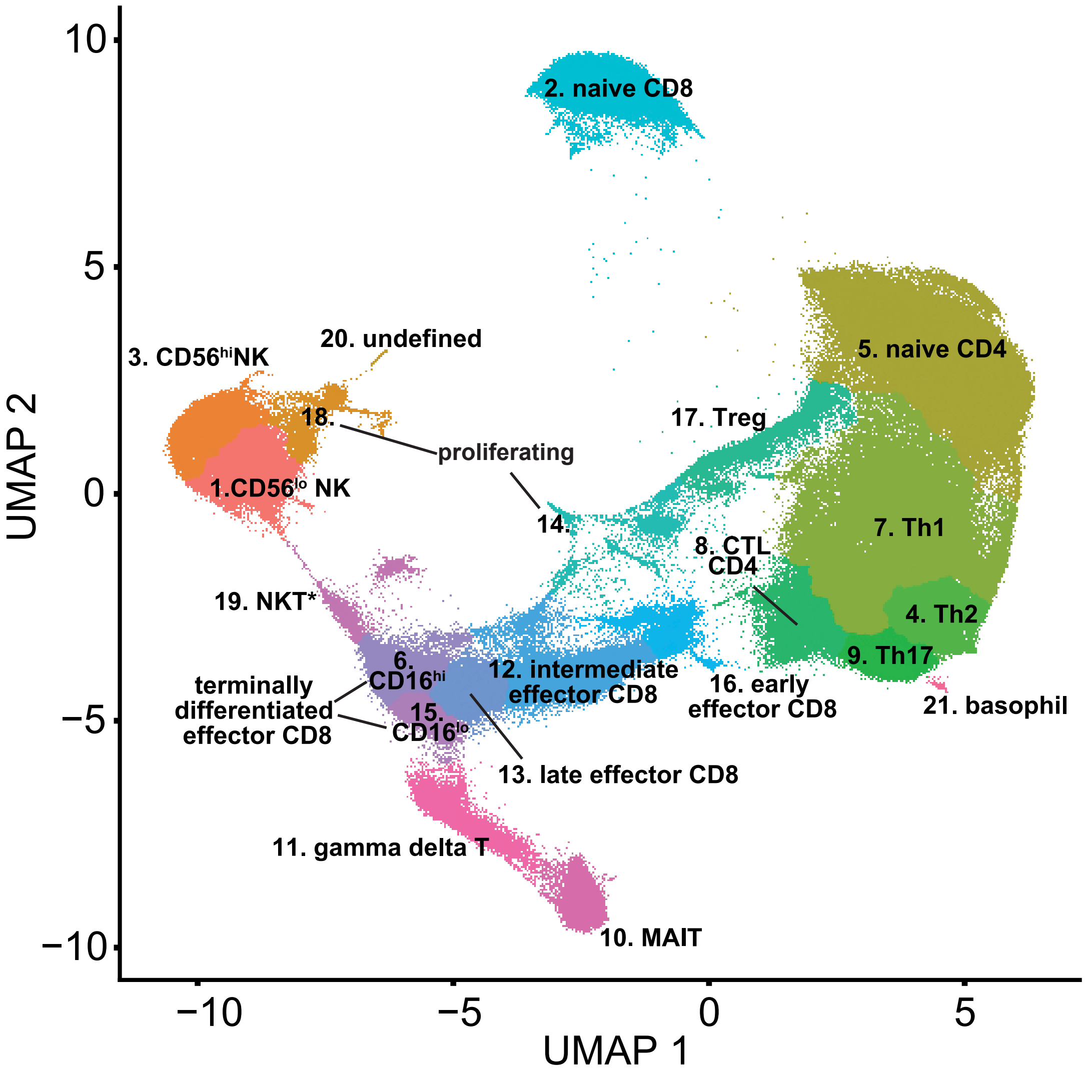

### Supplementary Fig. 7

#### Supplementary Figure 7

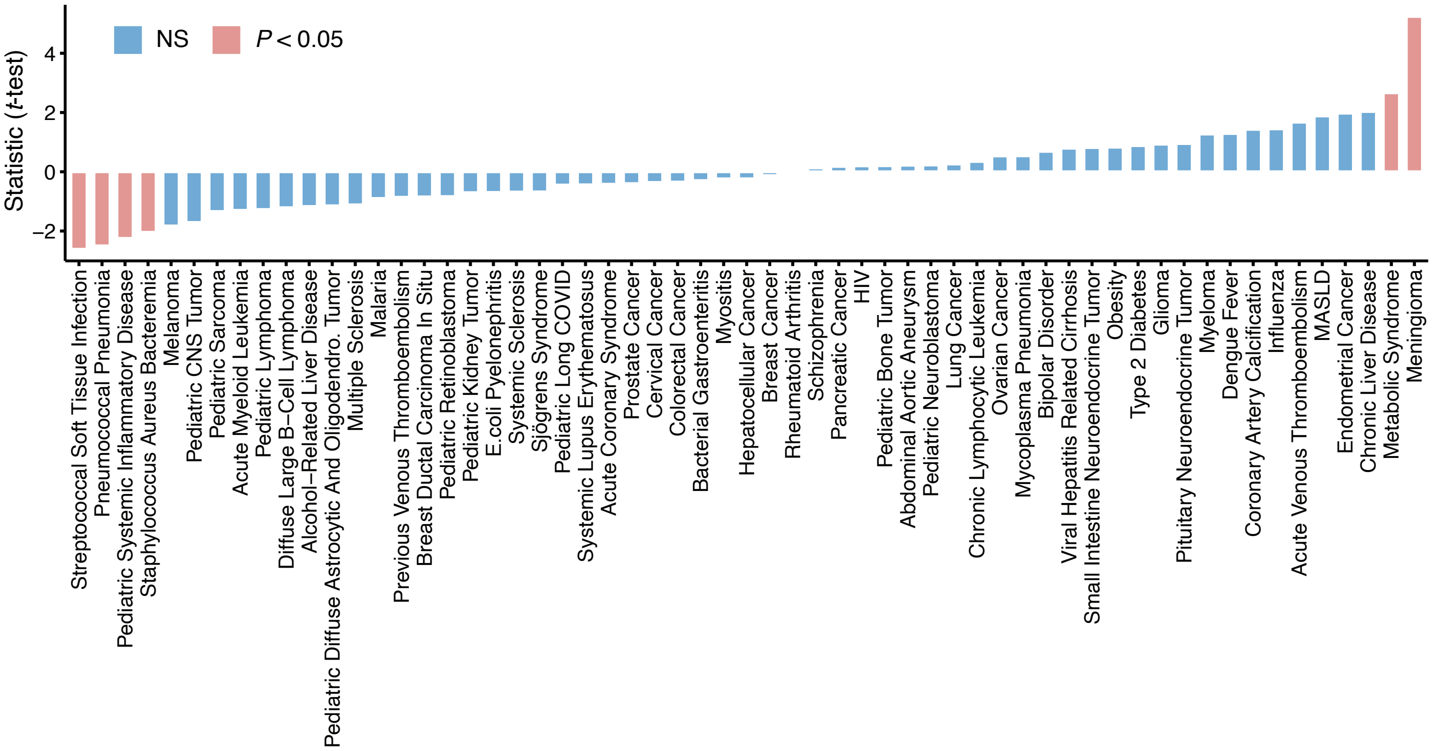
